# Identification of deleterious neutrophil states and altered granulopoiesis in sepsis

**DOI:** 10.1101/2022.03.22.22272723

**Authors:** Andrew J. Kwok, Alice Allcock, Ricardo C. Ferreira, Madeleine Smee, Eddie Cano-Gamez, Katie L. Burnham, Yasemin-Xiomara Zurke, Oxford acute medicine/ED research, Stuart McKechnie, Claudia Monaco, Irina Udalova, Charles J. Hinds, Emma E. Davenport, John A. Todd, Julian C. Knight

## Abstract

Sepsis is a condition of high mortality arising from dysregulation of the host immune response. Gene expression studies have identified multiple immune endotypes but gaps remain in our understanding of the underlying biology and heterogeneity. We used single-cell multi-omics to profile 272,993 cells across 48 whole blood samples from 26 sepsis patients (9 with paired convalescent samples), 6 healthy controls and 7 post-surgery patients. We identified immature neutrophil populations specific to sepsis and demonstrated the immunosuppressive nature of sepsis neutrophils in vitro. An *IL1R2*+ neutrophil state was expanded in a transcriptomic sepsis endotype associated with increased early mortality (sepsis response signature 1, SRS1), together with enrichment of the response to IL-1 pathway in mature neutrophils, marking IL-1 out as a potential target for immunotherapy in SRS1 sepsis patients. We confirmed the expansion of immature neutrophils, specifically *IL1R2+* neutrophils, in SRS1 in additional cohorts of patients (n = 906 RNA-sequencing samples, n = 41 CyTOF samples). Neutrophil changes persisted in convalescence, implicating disrupted granulopoiesis. Our findings establish a cellular immunological basis for transcriptomically defined sepsis endotypes and emphasise the relevance of granulopoietic dysfunction in sepsis, identifying opportunities for precision medicine approaches to the condition.

## Introduction

Sepsis describes a state of life-threatening organ dysfunction caused by a dysregulated host response to infection and is associated with a persistently high mortality of 20-30%(1). The axes of immune dysregulation have been examined by bulk transcriptomic studies(2–5) or single-cell interrogation of peripheral blood mononuclear cells (PBMCs)(6), with evidence of maladaptive inflammation and immunosuppression involving myeloid and lymphoid cells. However, there has been a lack of high resolution single-cell transcriptomic readouts inclusive of granulocytes, which form the majority of peripheral blood leukocytes, leading to a disconnect in interpreting single-cell PBMC vs. whole blood bulk transcriptomic data. As the first line of defense against pathogens, neutrophils have been demonstrated through peripheral blood immunophenotyping to be differentially abundant between sepsis and non-infectious inflammation(7). In sepsis patients, expansion of immunosuppressive granulocytes has been associated with a higher risk of nosocomial infection(8).

Individual variation in the sepsis response has been a major contributory factor in the failure of more than 100 randomised clinical trials of immunomodulatory therapies in sepsis(9). There is therefore a major need for better understanding of the nature of immune dysfunction in sepsis to enable precision medicine approaches e.g. targeted use of immunomodulatory therapies. We previously derived whole blood transcriptomic sepsis response signatures (SRSs) that were independent of source of infection(10), defining disease endotypes associated with early mortality(3, 10) and differential response to steroid therapy(11). Overlapping transcriptomic signatures with similar translational potential have been found by other groups(2, 4).

Recognising the need for mechanistic insight for an endotypic definition, we provide in the current manuscript evidence for the basis of the observed endotypes in sepsis and wider insights into disease pathophysiology. We sought to do this through an unbiased single cell multi-omic approach that considered all major immune cell types in peripheral blood including granulocytes. We elucidate the landscape of peripheral blood cellular immune alterations in sepsis, identifying a role for specific neutrophil populations in acute disease and persistent neutrophil changes in convalescence.

## Results

### Single-cell multi-omics recapitulates immune hallmarks of sepsis

We recruited a prospective cohort of 26 sepsis patients with 9 paired convalescent samples, 6 healthy volunteer (HV) controls, and 7 post-cardiac surgery (CS) samples as sterile inflammation controls for comparison to the sepsis patients (cohort 1). Our recruitment strategy encompassed sepsis patients admitted to the emergency department (ED) and intensive care unit (ICU). We screened patients admitted to the ED with suspected infection and symptoms and signs of established sepsis, with a change in quick SOFA score of ≥2 points(1). To enhance specificity, patients were recruited only if they also had a National Early Warning Score 2 (NEWS2), a physiological measure of acute illness severity, of 7 or above(12, 13) and were clinically adjudicated to have sepsis. Demographics and clinical details of the patients are detailed in Table S1.

To establish an unbiased single-cell atlas of peripheral leukocytes in sepsis patients, we profiled the whole-transcriptome and expression of 30 cell-surface proteins (Table S2) from fresh whole blood samples using the BD Rhapsody platform (Fig. 1A). A total of 272,993 high-quality single cells (Fig. S1) were retained for downstream analyses. We performed initial broad annotation of the dataset (Fig. 1B) by first manually annotating clusters based on lineage-specific protein marker expression (Fig. S2A), followed by cross referencing annotations with a data driven, algorithmic approach where annotations per cell were assigned based on reference RNA-seq profiles of immune cells from the Blueprint Consortium (Fig. S2B). We found the two approaches to be broadly concordant. At this level of annotation, we recapitulated well described hallmarks of peripheral blood immunophenotypes in sepsis including neutrophilia, lymphopenia and reduced classical monocyte (cMono) HLA-DR gene expression(14) (Fig. 1C).

**Figure 1.**
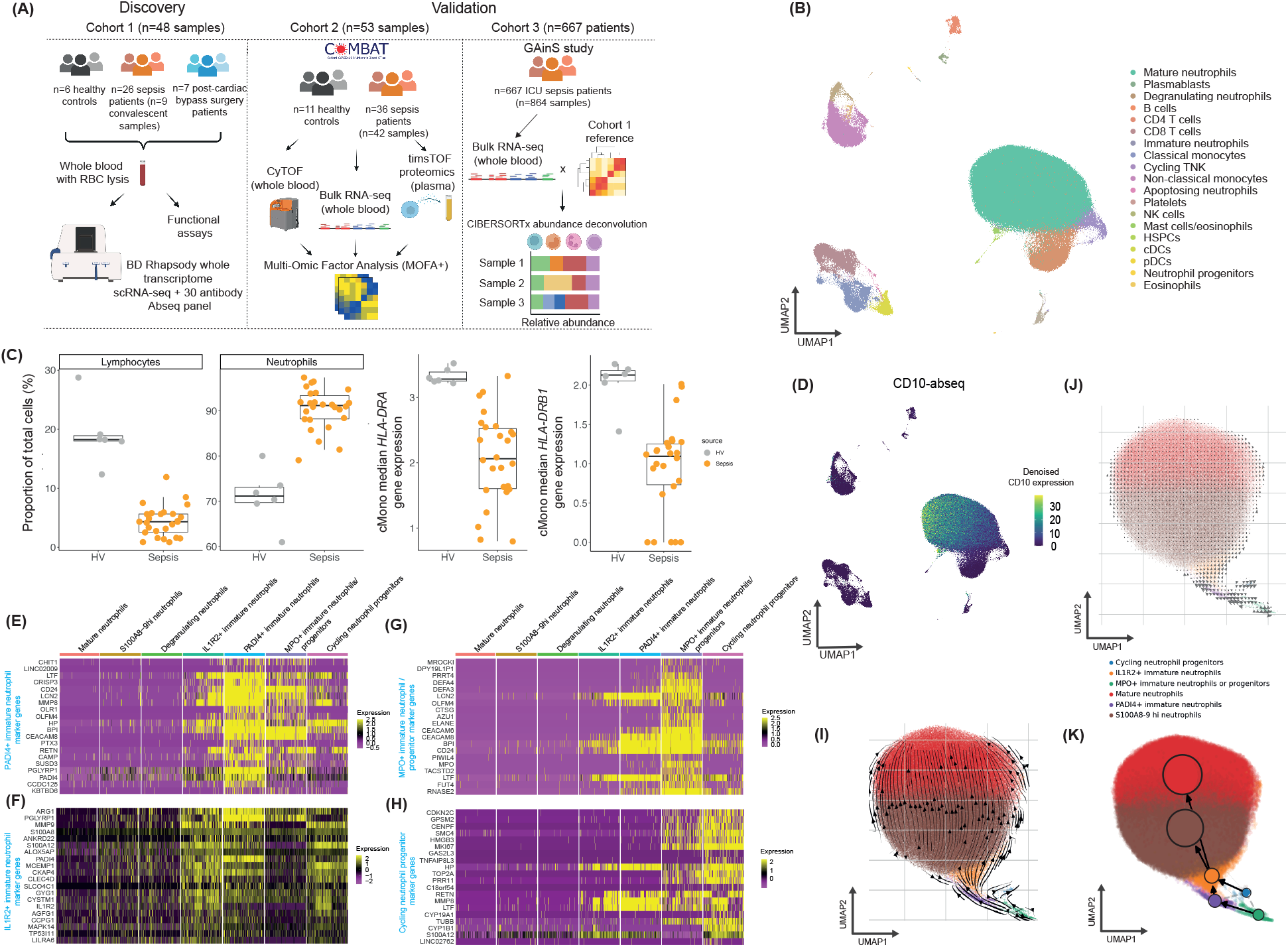
Unbiased peripheral blood total leukocyte single cell census in sepsis. (A) Schematic of experimental and analytical strategy for sepsis whole blood single cell immunoprofiling in a prospectively enrolled cohort (cohort 1) (n = 48 samples with 39 unique individuals) and validation cohorts (cohorts 2 and 3). (B) Uniform manifold approximation and projection (UMAP) visualisation of 272,993 cells after quality control, colored by broad annotations. (C) Boxplots of proportions of neutrophils and lymphocytes and *HLA-DRA* and *HLA-DRB1* gene expression in classical monocytes in sepsis vs. HV samples. (D) UMAP visualisation of all cells with denoised CD10 protein marker expression overlay. (E-H) Heatmaps of gene markers for (E) *PADI4+* immature neutrophils (F) *IL1R2+* immature neutrophils (G) *MPO+* immature neutrophils/progenitors and (H) cycling neutrophil progenitors. (I) UMAP visualisation of neutrophils (excluding degranulating neutrophils) with RNA velocity stream directions plotted. (J) UMAP visualisation as per (I) with size of velocity vector throughout vector field depicted. (K) Partition based graph abstraction analysis of RNA velocity predicted cellular trajectory transitions. *ICU = intensive care unit, TNK = T or NK cells, cMono = classical monocytes, cDCs = classical dendritic cells, pDCs = plasmacytoid dendritic cells, HSPCs = haematopoietic stem and progenitor cells*.

### Expansion of immature neutrophils in sepsis

To better characterise the role of specific cell states in our sepsis response atlas, we clustered cells at a higher resolution for fine annotation (Methods) (Fig. S2C, Table S3), noting the presence of multiple neutrophil clusters. CD10 expression (a marker of neutrophil maturity) visualised on uniform manifold approximation and projection (UMAP) suggested the presence of neutrophils at varying stages of maturation (Fig. 1D), in line with previous neutrophil single-cell analyses in other disease states(15, 16). Consistently, marker genes for these neutrophil clusters included multiple genes associated with immaturity, including *CD24* and *ARG1(*16) (Fig. 1E-F), as well as genes encoding neutrophil granule proteins such as *MPO, ELANE, AZU1, CTSG*, and *RNASE2* (Fig. 1G). We annotated the clusters according to gene markers that particularly distinguished the neutrophil state, including a cluster of neutrophils with marked *IL1R2* expression and a population undergoing cell cycle (Fig. 1H).

For analytical validation of the maturity annotations, we performed RNA velocity analysis on the neutrophil compartment to recapitulate differentiation trajectories based on splicing dynamics(17). Most velocity vectors started from clusters annotated as progenitors (Fig. 1I), forming a single overall trajectory. The magnitude of the velocity vector field was substantially greater in the areas of the immature neutrophil clusters, suggesting that most of the differentiation stopped once either the *S100A8/9* high neutrophil state or mature neutrophil state were reached (Fig. 1J). Partition based graph abstraction analysis(18) supported a sequential progression of maturation according to the observed clusters into two terminal states of differentiation i.e. the annotated mature neutrophils and *S100A8/9* high neutrophils (Fig. 1K).

We next investigated the cell compositional changes in sepsis at the fine level of annotation (Fig. 2A). Sampling neighborhoods of cells from the *k*-nearest neighbors graph used for clustering (Methods) (19), we examined differential abundance across conditions by looking for enrichment of either condition in each neighborhood. Neighborhoods in multiple cell states differed significantly in proportion, with all immature neutrophil populations, as well as degranulating and *S100A8/9* high neutrophils, increased in sepsis compared to HV (Fig. 2B), while all mononuclear cell (MNC) subsets except plasmablasts were reduced. A similar pattern of compositional remodeling could be observed when comparing CS patients to HVs, suggesting that the changes detected reflected non-specific features of inflammation (Fig. S3A).

**Figure 2.**
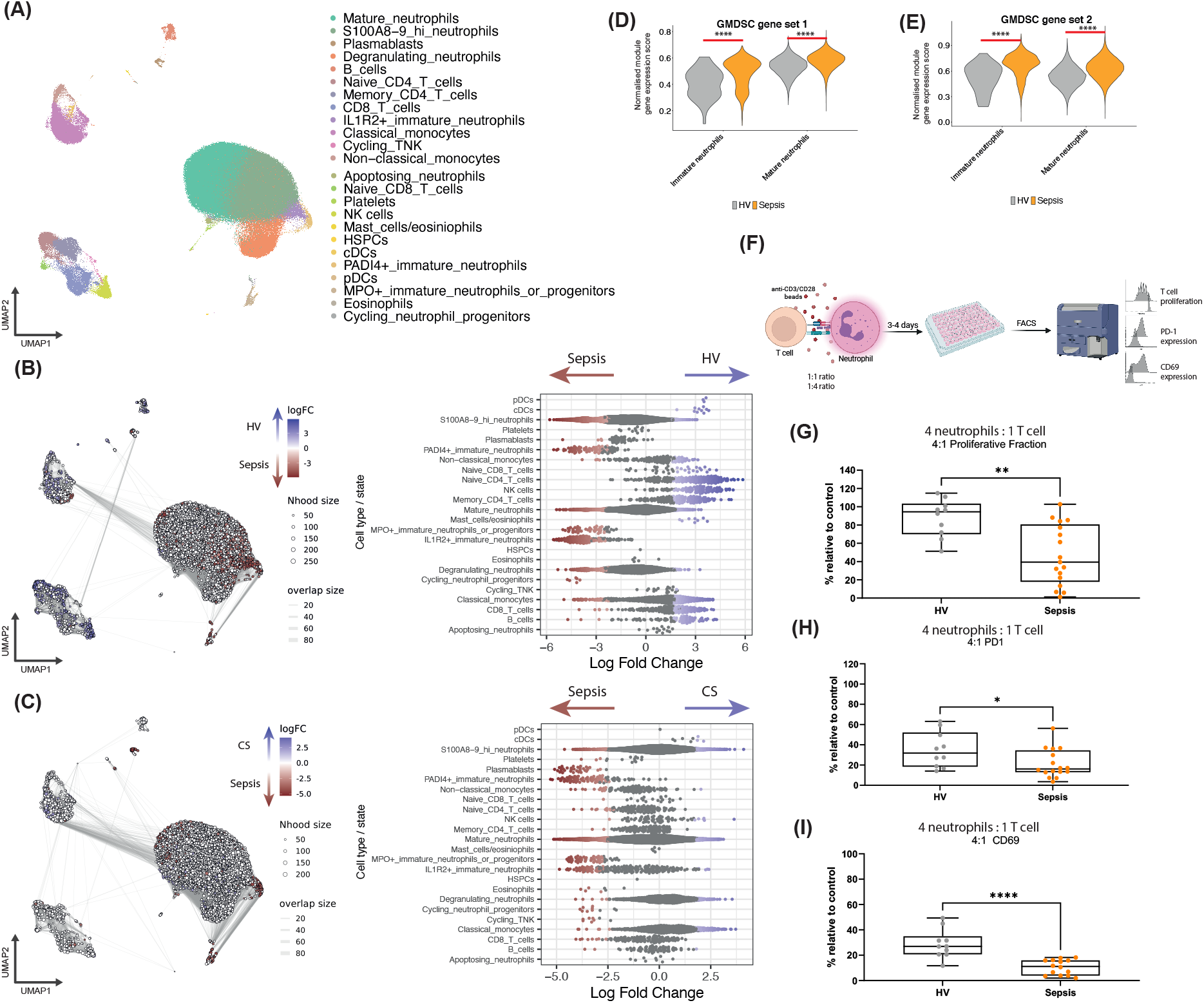
Emergence of immature neutrophils in sepsis with an immunosuppressive function. (A) UMAP visualisation of all cells colored by fine annotations. (B) Differential abundance (DA) analysis using graph neighborhood-based method to detect enrichment of neighborhoods per comparator condition between sepsis and HV samples, with UMAP visualisation of sampled neighborhoods and statistically significant enrichment colored (spatial FDR < 0.05 with generalised linear modelling) (red = enrichment in sepsis, blue = enrichment in HV) and corresponding beeswarm plot (right) with cluster labels of neighborhoods depicted. (C) DA analysis between sepsis and CS samples. (D-E) Gene set scoring of mature and immature neutrophils in sepsis and HV samples by gene signatures of granulocytic myeloid derived suppressor cells (two-sided Wilcoxon rank-sum test). (F) Schematic of 3-4 day neutrophil-allogeneic CD4 T cell co-culture experiment to detect immunosuppressive properties of neutrophils. (G-I) Boxplots of proliferative fraction (G), surface PD-1 positive cells (H) and surface CD69 positive cells (I) of anti-CD3/28 bead stimulated CD4 T cells after co-culture with neutrophils at a 4 neutrophil : 1 T cell ratio, compared to positive controls of CD4 T cells cultured with anti-CD3/28 beads alone (two-sided Wilcoxon rank-sum test). \**P* < 0.05, \*\**P* < 0.01, \*\*\*\**P* < 0.0001. *cDCs = classical dendritic cells, pDCs = plasmacytoid dendritic cells, HSPCs = haematopoietic stem and progenitor cells, FC = fold change, FDR = false discovery rate, GMDSC = granulocytic myeloid derived suppressor cells, HV = healthy volunteers, CS = cardiac surgery patients*.

When contrasting sepsis patients with CS patients, we found significant enrichment of the four immature neutrophil populations in sepsis (Fig. 2C). Thus, whereas the commonly recognised hallmarks of sepsis (neutrophilia, lymphopenia(20) and low cMono HLA-DR(14)) may in fact be more reflective of a non-specific inflammatory insult (Fig. S3B), immature neutrophils appeared to be specific to sepsis.

### Sepsis neutrophils show CD4 T cell immunosuppressive capacity

Immature neutrophils have frequently been suggested to be immunosuppressive but many reports that tie immaturity to immunosuppression rely purely on phenotypic observation. While upregulation of immunosuppressive pathways was not apparent from differential gene expression analysis (Fig. S3C-D, Table S4), on scoring our neutrophil populations for expression of published gene sets derived from experimentally validated immunosuppressive neutrophils (granulocytic myeloid derived suppressor cells, G-MDSCs)(21, 22), we found a significantly higher enrichment score of these gene sets in both mature and immature neutrophils from septic patients compared to those from HVs (Wilcoxon rank sum tests, FDR < 0.0001) (Fig. 2D-E).

To further test the immunosuppressive properties of sepsis neutrophils, we designed an *in vitro* neutrophil-allogeneic CD4 T cell co-culture system (Fig. 2F). We found that upon culturing CD4 T cells with neutrophils in the presence of anti CD3/28 beads, sepsis neutrophils significantly inhibited CD4 T cell proliferation (measured by fraction of proliferating T cells) and activation (measured by percentage of T cells positive for PD-1 and CD69 expression) (Fig. 2G-I), with a dose response effect (Fig. S4A-C). Increased arginase-1 activity and PD-L1/2 inhibition of T cells have been proposed as mechanisms for neutrophil immunosuppression(7, 8), but we were unable to reverse the suppressive effects with either anti-PDL1/2 antibodies or arginase-1 inhibitors (Fig. S4D). Instead, we were able to partially reverse suppression of T cell activation by inhibiting the prostaglandin EP2 receptor, as reflected by partial restoration of CD4 T cell CD69 expression in sepsis neutrophil co-cultures when adding the EP2 receptor inhibitor TG6-10-1 (Fig. S4D). Neither addition of indomethacin (cyclo-oxygenase inhibitor) nor inhibition of the prostaglandin EP4 receptor showed a similar effect, suggesting a mechanism of action at the level of pre-formed prostaglandin E2 acting at the EP2 receptor (Fig. S4D). Moreover, we were only able to partially restore CD69 expression and could not modulate PD-1 expression or proliferation suppression, suggesting that other mechanisms explain the neutrophil mediated CD4 T cell suppression.

In contrast to previous reports(7), when we subjected the neutrophils to other functional tests, we found that cell-intrinsic functions of sepsis neutrophils were intact, with preserved phagocytic activity and ROS production (Figs. S4E-F).

### *IL1R2*+ immature neutrophils underlie a deleterious sepsis endotype

Beyond resolving the differences in cell states between sepsis and sterile inflammation or health, heterogeneity within sepsis remains a major challenge. A major clinical question is how to employ methods to stratify patients for meaningful prognostication and targeted treatments. Understanding the molecular drivers of patient subgroups could inform rational approaches to therapeutic trials. To this end, we classified patients in our cohort based on our previously proposed Sepsis Response Signatures (SRS), which were defined using peripheral blood bulk transcriptomics and robustly replicated across cohorts and causes of sepsis(3, 10). The SRS signature can be described using discrete patient endotypes, of which SRS1 is known to exhibit higher early mortality, as well as using a quantitative score (SRSq) known to positively correlate with the degree of immune dysfunction and illness severity(23). SRS1 patients and patients with high SRSq score display features of immunosuppression.

We assigned SRS status and SRSq scores to our 26 acute sepsis samples using the previously defined seven gene set (*DYRK2, CCNB1IP1, TDRD9, ZAP70, ARL14EP, MDC1*, and *ADGRE3*) and the algorithm implemented in the SepstratifieR package(23). Based on pseudobulk profiles of our whole blood single-cell dataset, we identified 16 SRS1 samples and 10 non-SRS1 samples. Principal components analysis (PCA) of the top 15% most variable genes of our pseudobulk profiles showed separation of the SRS groups along the first two principal components (PCs) (Fig. S5A). The robustness of the SRS assignment was assessed by correlating the fold changes in gene expression between SRS1 vs. non-SRS1 samples with the fold changes in the cohort from which SRS was originally derived(3). We found a strong and highly significant correlation, supporting the validity of SRS signals in this cohort (Fig. S5B).

Testing for cellular differential abundance across SRS groups revealed an expansion of *IL1R2*+ immature neutrophils in SRS1 compared to non-SRS1 patients, as well as minor enrichments in *S100A8/9* high neutrophils, degranulating neutrophils and cycling neutrophil progenitors, whereas in non-SRS1 patients there was enrichment of NK cells, memory CD4 and CD8 T cells and cMonos (Fig. 3A).

**Figure 3.**
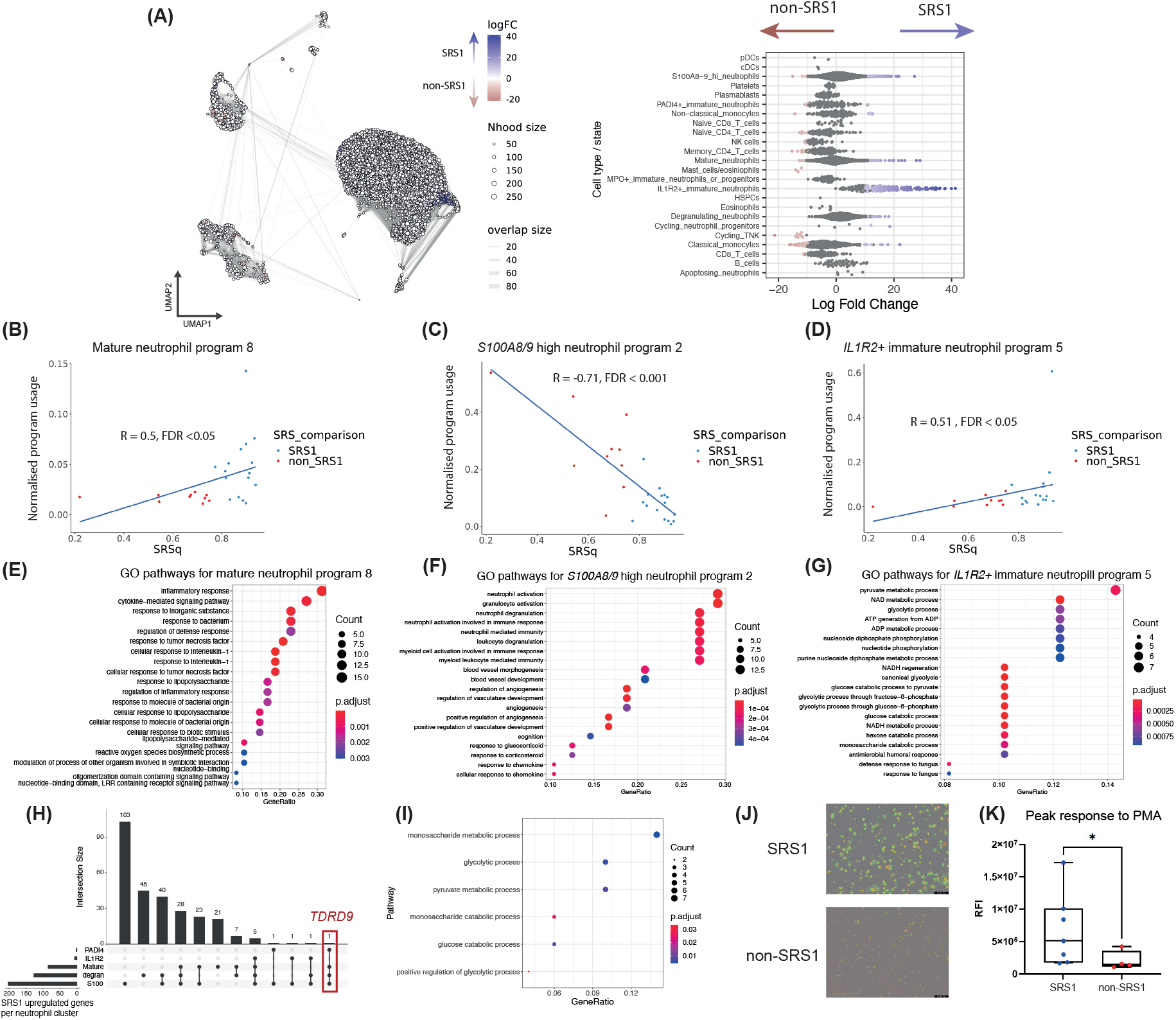
Immature IL1R2+ neutrophils and neutrophil specific DGE drive a deleterious septic state (SRS1). (A) Differential abundance (DA) analysis using graph neighborhood-based method to detect enrichment of neighborhoods per comparator condition between SRS1 and non-SRS1 samples, with UMAP visualisation of sampled neighborhoods and statistically significant enrichment colored (spatial FDR < 0.05 with generalised linear modelling) (red = enrichment in non-SRS1, blue = enrichment in SRS1) and corresponding beeswarm plot (right) with cluster labels of neighborhoods depicted. (B-D) Representative correlation plots of gene programs with program expression significantly correlated (Spearman *R* FDR < 0.05) with SRSq for mature neutrophils (B, program 8), *S100A8/9* high neutrophils (C, program 2), and *IL1R2+* immature neutrophils (D, program 5). (E-G) Over-representation test pathway enrichment dotplots of top Gene Ontology (GO) pathways for gene programs of (E) mature neutrophils (program 8) (F) *S100A8/9* high neutrophils (program 2) and (G) *IL1R2+* immature neutrophils (program 5). (H) UpSet plot of overlap in differentially expressed genes between SRS1 and non-SRS1 across different neutrophil subsets. (I) Over-representation test pathway enrichment dotplot of GO glycolysis related gene sets for top 0.05% genes correlated with *TDRD9* expression in all neutrophils. (J) Representative images of neutrophils undergoing NETosis from a SRS1 patient vs. a non-SRS1 patient at 4 hours post PMA stimulation, where fluorescence green from Cytotox green dye denotes cell actively undergoing NETosis. (K) Boxplot of peak Cytotox green fluorescence intensity per sample by SRS group (two-sided Wilcoxon rank sum test). \**P* < 0.05. *SRS = sepsis response signature, FC = fold change, cDCs = classical dendritic cells, pDCs = plasmacytoid dendritic cells, HSPCs = haematopoietic stem and progenitor cells, FDR = false discovery rate, PMA = phorbol 12-myrisate 13 acetate. RFI = relative fluorescence intensity*.

To understand the cellular basis of SRS, we performed pseudobulk differential gene expression (DGE) analysis separately for each cell state, testing against SRS as the covariate of interest, and adjusting for age, sex and sequencing batch. Strikingly, we found minimal DGE in any of the MNC subsets (Fig. S5C), suggesting a minor contribution of MNCs to SRS definitions. By contrast, we found substantial numbers of differentially expressed genes (DEGs) at a FC of 1.5 and FDR < 0.05 for mature neutrophils, *S100A8/9* high neutrophils, degranulating neutrophils, *IL1R2*+ immature neutrophils and *PADI4*+ neutrophils (Fig. S6A), which translated into separation of samples by SRS status on PCA (Fig. S6B), suggesting that gene expression profiles in neutrophils were the underlying drivers of SRS (Table S5).

As genes function in concert in biological pathways and programs, we conducted consensus non-negative matrix factorisation (cNMF)(24) of each of the neutrophil states to further elucidate differential activity between SRS state at the level of gene programs rather than of individual genes (Tables S6-10). Statistical testing of program activity revealed that at FDR < 0.05 three programs were correlated with SRSq scores in mature neutrophils and *S100A8/9* high neutrophils, four in *IL1R2*+ neutrophils, and two in *PADI4*+ immature neutrophils (Fig. 3B-D and Fig. S7A-D). Over-representation enrichment testing of the top 50 genes of each of these differentially active programs revealed (Fig. S7A-D): 1) pathways including response to IL-1 for mature neutrophils (Fig. 3E), notable given the expansion of *IL1R2+* neutrophils, as *IL1R2* is a decoy receptor for IL-1β; 2) response to glucocorticoids in *S100A8/9* high neutrophils (Fig. 3F), notable given the interaction between steroid administration, SRS and mortality in sepsis(11); and 3) the glycolytic pathway in *IL1R2*+ immature neutrophils (Fig. 3G), notable given the shifts to aerobic glycolytic metabolism previously noted in sepsis(25).

We found that *TDRD9*, one of the genes used to classify SRS state, was strongly upregulated in four of six neutrophil states (notably not in *IL1R2+* immature neutrophils) (Fig. 3H). To identify potentially co-regulated genes associated with SRS status, we analysed *TDRD9* co-expression by ranking all genes according to their correlation with *TDRD9* expression in all neutrophils. Pathway analysis of the top 0.5% of genes in this guilt by association ranking returned enrichment in glycolysis among other pathways, suggesting that the shift in metabolic state observed in SRS1 was present not only in *IL1R2*+ immature neutrophils but rather was a more general phenomenon in SRS1 neutrophils (Fig. 3I).

Functional testing of neutrophils between SRS states did not show differences in neutrophil CD4 immunosuppressive potential or ROS production (Figs. S8A-D). However, we found that SRS1 neutrophils displayed reduced phagocytosis compared to non-SRS1 neutrophils, in line with previous observations of greater immunosuppression in SRS1 (Fig. S8E). Given the glycolytic shift we identified, we hypothesised a potential link to NETosis, another cell-intrinsic function of neutrophils that is known to be dependent upon glycolysis. Consistent with this hypothesis, we found SRS1 neutrophils exhibited greater NETosis than non-SRS1 neutrophils upon stimulation with phorbol 12-myrisate 13 acetate (PMA) (Fig. 3J-K).

### Validation of neutrophil immaturity in SRS1 in further sepsis cohorts

To test the validity of our results, we first sought further evidence of increased immature neutrophils in SRS1. We re-analysed whole blood bulk RNA-seq and mass cytometric (CyTOF) immunophenotyping data from a cohort of 36 all cause sepsis patients (42 samples), who were previously recruited to the same study and included as part of a bioresource from the COMBAT consortium to compare with COVID-19 patient samples (cohort 2)(26) (Fig. 1A, Table S11). We assigned SRS endotypes in this cohort; PCA of the top 15% most variable genes showed separation of the two SRS groups along PC1 and PC2 (Fig. S9A). The overall 14-day mortality in this cohort was 25%, with SRS1 patients having a significantly higher early mortality (14-day mortality log-rank test *P* = 0.0045, 42.8% versus 0%) (Fig. 4A), emphasising the relevance of SRS endotypes for the clinical stratification of sepsis patients.

**Figure 4.**
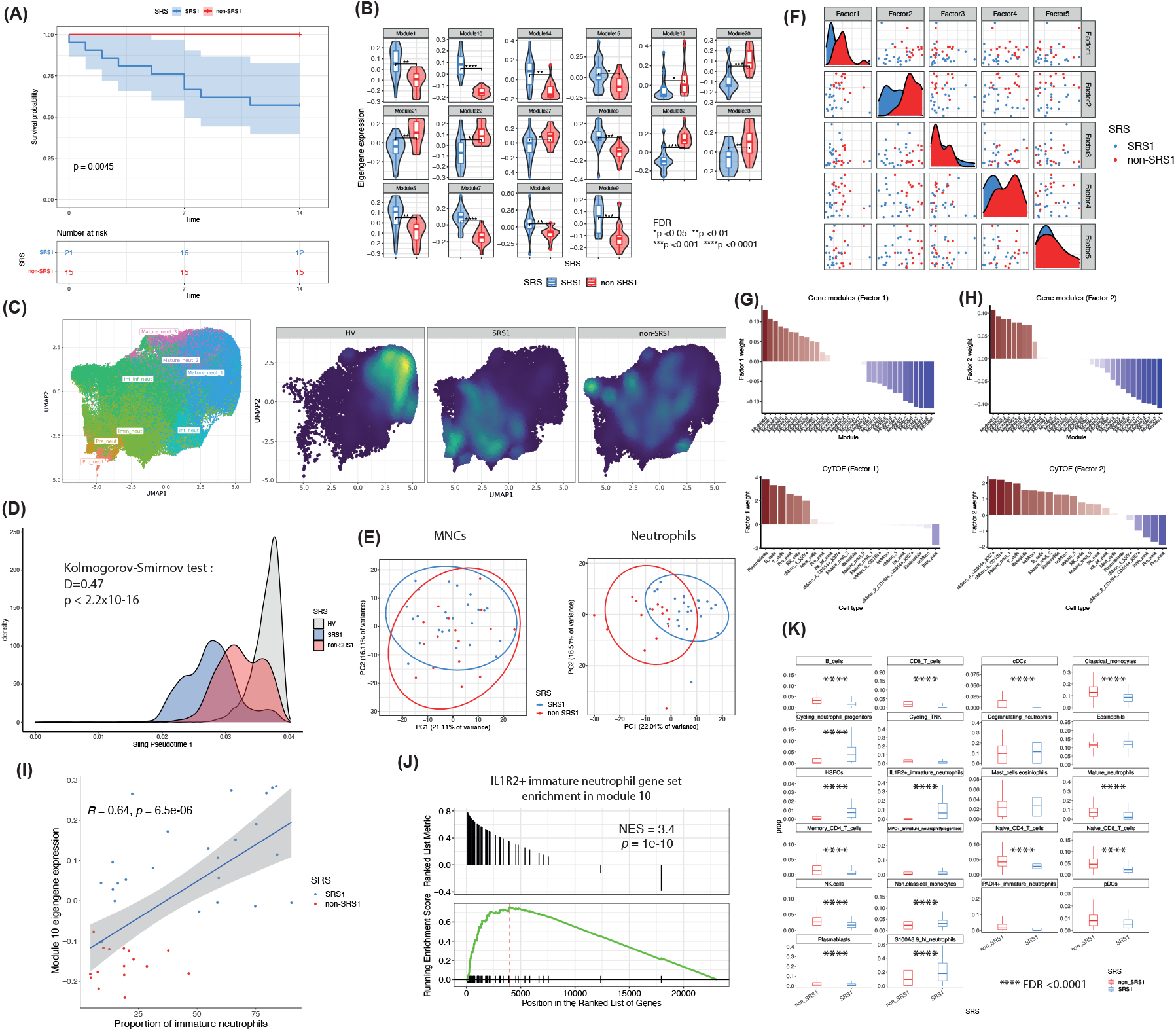
Validation of neutrophil immaturity underlying SRS1 and *IL1R2*+ immature neutrophil expansion in SRS1. (A) Kaplan-Meier survival curves of days after research blood sampling by SRS groups (shaded areas depict 95% confidence intervals) with *P-*value from log rank test. SRS grouping was taken using the latest sample for patients with multiple samples. (B) Violin and boxplots of 16 differentially expressed module eigengenes between SRS1 and non-SRS1 with FDR shown (two-sided Wilcoxon rank sum test). (C) UMAP visualisation of neutrophil subsets (1,921,471 cells) and density-based visualisation of neutrophil distributions faceted by SRS (and healthy volunteer [HV]) status. (D) Density distribution of neutrophils over pseudotime. (E) First two principal components from principal components analysis of mononuclear cell proportions and neutrophil proportions colored by SRS status. (F) Scatter and density plots of factor values for latent factors one to five, derived from factor analysis of 33 gene modules, 22 cell types and 105 plasma proteins for n=36 sepsis patients, colored by SRS group membership. (G-H) Barplots of weights per gene module and cell type for (G) latent factor 1 and (H) latent factor 2 colored by weight (red-blue gradient for positive to negative weights). (I) Scatterplot of module 10 eigengene expression against proportion of immature neutrophils per sample of cohort 2 colored by SRS assignment, with Pearson *R* correlation. (J) Leading edge analyses for enrichment of *IL1R2+* immature neutrophil defining signature in module 10. (K) Boxplots of cell type/state proportions in n=667 intensive care unit SRS assigned sepsis samples derived from bulk RNA-seq data deconvolution by CIBERSORTx with reference matrix built by cohort 1 single cell RNA-seq data (two-sided Wilcoxon rank sum test). *FDR < 0.05, **FDR < 0.01, *** FDR < 0.001, **** FDR < 0.0001. *SRS = sepsis response signature, FDR = false discovery rate, MNCs = mononuclear cells, NES = normalised enrichment score*.

We reduced dimensions of the RNA-seq dataset with weighted gene co-expression network analysis (WGCNA)(27) (Methods). Of the 33 distinct gene modules extracted, differential eigengene expression testing for the 36 individuals yielded nine upregulated and seven downregulated modules in SRS1 (Wilcoxon rank sum test FDR < 0.05) (Fig. 4B). Overlapping the module genes with signatures of differentiating and differentiated neutrophils from mouse single cell transcriptomics (28), we found that modules 7, 10 and 14 were significantly enriched for differentiating neutrophils (Fig. S9B).

To elucidate the cellular basis of these signals in SRS1, we re-analysed CyTOF data available for 41 of the 42 sepsis samples in COMBAT and 11 age and sex matched healthy controls by iterative clustering, merging and annotating cell types and states (Fig. S9C). We subclustered the neutrophils with 17 selected markers, assigning them to eight distinctively different states according to Marker Enrichment Modelling(29) which spanned a range of maturity stages (Fig. 4C and Fig. S9D). This was confirmed by pseudotime trajectory analysis with diffusion map dimensionality reduction and principal curve fit *(30)* (Fig. S9E-F), where trajectory one ordered cells along pseudotime (Fig. S9G).

We found that the density of neutrophils in the earlier and more immature stages of the trajectory was significantly greater in SRS1 than non-SRS1 (Fig. 4D, Kolmogorov-Smirnov test D=0.47, *P* < 2.2×10^−16^). Density visualisation on UMAP of neutrophil subsets illustrated the skewing of neutrophil populations to either pro-, pre-neutrophil and immature phenotypes in SRS1 vs. non-SRS1 (Fig. 4C), which was confirmed as significant differential abundance with mixed models (Fig. S9H). PCA on cell proportions within the neutrophil and MNC compartments showed that, in contrast to neutrophils, proportional differences in MNCs in isolation could not separate the SRS groups (Fig. 4E).

To understand the relationship between the differences in cell composition and the differences in bulk gene expression profiles across SRS, we conducted factor analysis using MOFA+ to identify latent spaces containing shared variation across the two data modalities which would directly implicate the cell types driving the SRS gene expression signatures(31). We included additional data for the levels of 105 plasma proteins measured by mass spectrometry for 30 of the 36 individuals as a negative control (Fig. S9I), reasoning that the latent factors linking cell subsets to gene expression profiles should not show an overly strong contribution from the plasma proteomic data. Factors one and two were responsible for most of the observed SRS separation (Fig. 4F) and were significantly different between SRS groups (Fig. S9J). These two factors explained minimal variance within the plasma proteomic dataset. By contrast, factor five, which accounted for most of the variance in the plasma proteomic data (20.3%), was not different across SRS and accounted for < 0.01% of variance in the gene expression and CyTOF data (Fig. S9K). This suggested that differences in plasma protein levels did not contribute substantially to SRS, while the genes and cell types separating SRS1 and 2 along the first two factors were genuine drivers of SRS.

Of the 13 WGCNA gene modules driving a low factor one value (in the direction of SRS1), seven (modules 3, 5, 8, 7, 9, 10 and 1) were significantly upregulated in SRS1 in the differential co-expression analysis (Fig. 4B and 4G). Conversely, four of seven modules (modules 32, 33, 27 and 22) contributing to a high factor one value (in the direction of non-SRS1) were significantly upregulated in non-SRS1 (Fig. 4B and Fig. 4G). The key cell type contributing to the SRS1 direction was immature neutrophils, while the various MNCs (T, B and NK cells and plasmablasts) contributed in the opposite direction (Fig. 4G). Similarly, six of the top eight modules contributing to a negative factor two value in the direction of SRS1 (modules 1, 7, 14, 15, 10 and 9; Fig. 4H) were upregulated in SRS1 (Fig. 4B), while all five of the top modules leading to a positive factor two value had been significantly upregulated in non-SRS1. Correspondingly, we found strong negative weighting in the CyTOF modality for factor two to include pre-neutrophils, pro-neutrophils and immature neutrophils (Fig. 4H), highlighting the relevance of these immature neutrophil populations to the SRS1 state.

### Validation of *IL1R2+* neutrophil expansion in SRS1

While the analysis for cohort 2 revealed expansion of immature and progenitor neutrophil states, we aimed to directly test for enrichment of the *IL1R2*+ immature neutrophil population observed in cohort 1 in this second cohort. We identified module 10 as the gene module most correlated with the immature neutrophils as defined by CyTOF (Pearson *R* = 0.64, *P* = 6.5×10^−6^) (Fig. 4I). Leading edge analysis of genes in this module ranked by correlation with eigengene expression indeed showed enrichment of a signature defining *IL1R2+* immature neutrophils (normalised enrichment score [NES] 3.4, *P* = 1×10^−10^) (Fig. 4J), providing direct evidence of the concordance of our analysis between the two datasets.

To further validate these observations, we performed cell type/state proportion deconvolution with CIBERSORTx(32) on 864 SRS-assigned sepsis samples from the GAinS study for which only bulk whole blood gene expression data were available (Fig. 1A). This cohort consisted of adult sepsis patients recruited from the intensive care unit (ICU) with samples collected within 5 days of ICU admission. Using our single-cell multi-omics dataset as a reference, differential abundances of cell populations predicted from deconvolution for the 667 first available samples per patient showed significant MNC depletion and *IL1R2*+ immature neutrophil and cycling neutrophil progenitor expansion in SRS1 using univariate tests (Fig. 4K), recapitulating the observations from our multi-modal analyses.

### Persistent alterations to granulocyte compartment in convalescence

The appearance and expansion of various immature neutrophil subsets in sepsis provided circumstantial evidence for disrupted myelo/granulopoiesis (Fig. 2B-C). Univariate testing for differential plasma cytokine abundance of the myelopoietic cytokines GM-CSF, G-CSF and M-CSF in cohort 2 revealed that levels of all three cytokines were significantly higher in sepsis compared to HV samples (Fig. S10A). However, only G-CSF was found in significantly higher levels in SRS1 than in non-SRS1 (Fig. 5A), suggesting a specific granulopoietic skew to the myelopoiesis in SRS1.

**Figure 5.**
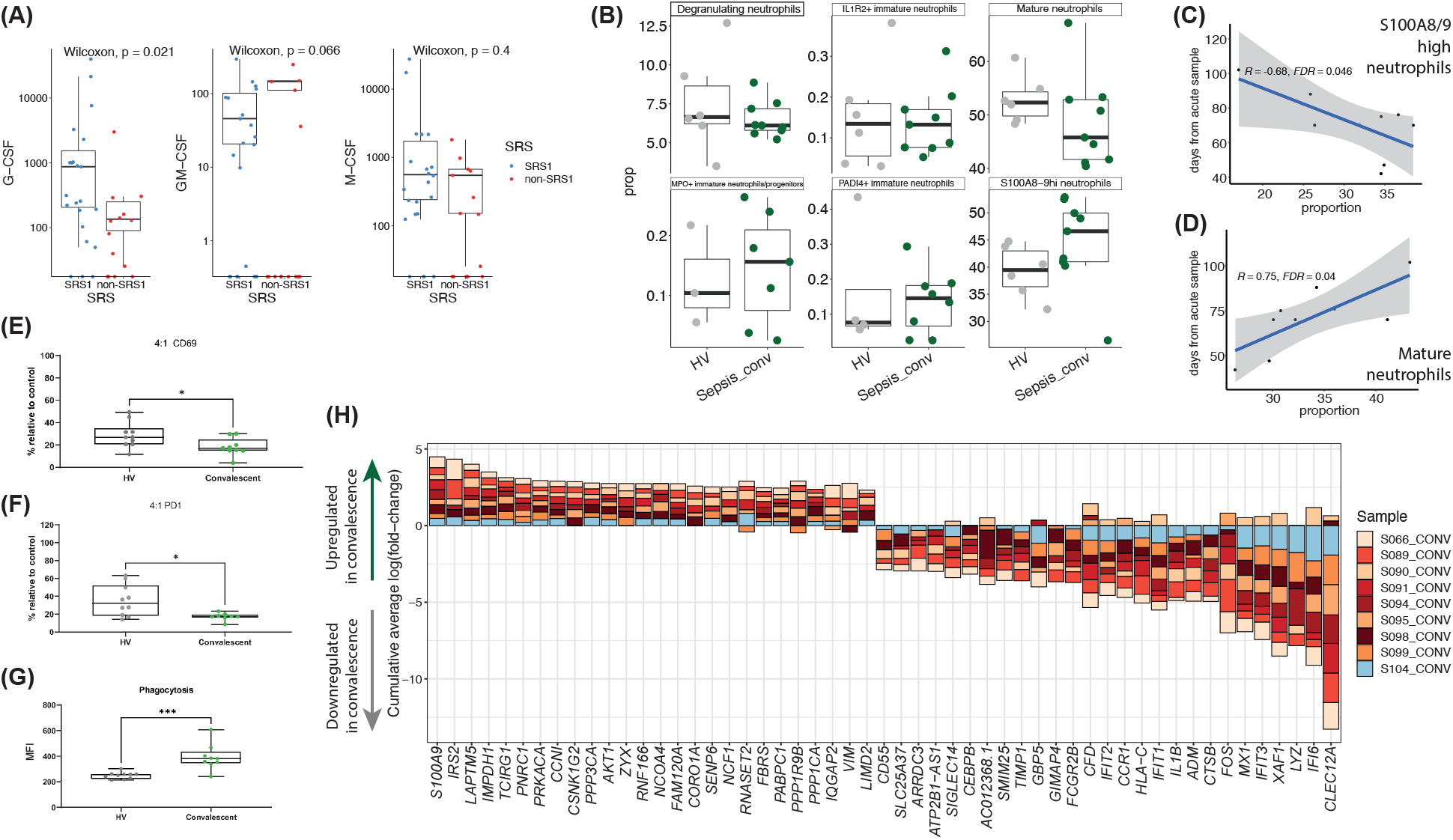
Evidence for disrupted granulopoiesis: persistent granulocytic compartment changes in convalescence. (A) Boxplots of plasma myelopoietic cytokine quantities between SRS groups in cohort 2 (two-sided Wilcoxon rank sum test). (B) Boxplots of proportions of neutrophil states in HV vs. convalescent sepsis samples from cohort 1. (C-D) Correlation (Pearson *R*, FDR < 0.05) of days from acute sample (for convalescent sample) against proportion of *S100A8/9* high neutrophils (C) and mature neutrophils (D). (E-F) Boxplots of surface CD69 positive cells (E) and surface PD-1 positive cells (F) of anti-CD3/28 bead stimulated CD4 T cells co-cultured with neutrophils, compared to positive controls of CD4 T cells cultured with anti-CD3/28 beads alone (two-sided Wilcoxon rank sum test). (G) Boxplot of median fluorescence intensity (MFI) of neutrophils for phagocytosis assay with fluorescently labelled *E. Coli* conjugated phagocytosis beads (two-sided Wilcoxon rank sum test). (H) Consensus differential gene expression analysis stacked barplots per convalescent sample neutrophils against HV neutrophils. *FDR < 0.05, **FDR < 0.01, *** FDR < 0.001. *SRS = sepsis response signature, FDR = false discovery rate, Sepsis_conv = convalescent sepsis*.

For a considerable time post hospital discharge, sepsis survivors are at high risk of readmission to hospital, most often due to recurrent infection(33). This clinical observation could be due to persistent changes in the innate immune system, including potential disrupted granulopoiesis. To test the hypothesis of granulopoietic disturbance in sepsis beyond endotypes, we analysed nine convalescent sepsis samples from cohort 1 collected one to three months after hospital discharge. We reasoned that, given the short lifespan of neutrophils, any changes observed in these samples must be attributable to altered granulopoiesis(34).

When comparing cellular composition between HVs and convalescent sepsis samples, we found no significant differences, although there were trends toward fewer mature neutrophils and more *S100A8/9* high neutrophils in convalescent samples (Fig. 5B). We examined the correlation between the proportion of these two neutrophil populations and time from acute sampling, finding strong and significant correlations for both populations (*R* = −0.68 FDR = 0.046 for S100A8/9 high neutrophils, *R* = 0.75 FDR = 0.04 for mature neutrophils), pointing to a recovery process whereby the proportion of *S100A8/9* high neutrophils decreases as sepsis resolves, with gradual resumption of mature neutrophil production. Functionally, this manifested as a persistent partial immunosuppression of CD4 T cells: whereas the suppression of CD4 T cell proliferation was no longer observed with convalescent sepsis neutrophil cultures (Fig. S10B), CD4 T cell CD69/PD-1 expression was still significantly reduced (Fig. 5E-F).

Although there was no difference in phagocytic ability between septic and HV neutrophils in cohort 1 (Fig. S4E), we found a significant increase in phagocytosis in convalescent sepsis neutrophils compared to HV neutrophils (Fig. 5G). No differences were observed for convalescent neutrophil ROS production or NETosis (Fig. S10C-D). Taken together, our functional data reflected elements of both trained immunity and continued immunosuppressive potential in neutrophils produced in convalescence, directly implicating altered granulopoiesis in the longer-term immune consequences of sepsis.

Finally, we examined gene expression changes in the convalescent sepsis samples more closely. Consensus DGE of total neutrophils between the convalescent sepsis and HV samples identified a number of genes with defense function downregulated in convalescence (Fig. 5H), including *IL1B* and type I interferon pathway genes such as *IFIT1, IFIT2, IFIT3, IFI6 and MX1*. These observations suggested that the neutrophil compartment had yet to return to the healthy state completely, and together with the persistent CD4 T cell suppressive changes, may provide the functional basis for patients’ continued susceptibility to infection and sepsis after recovery from an index episode.

## Discussion

In this study, we extend established hallmark immunophenotypes of sepsis and identify the specificity of immature and progenitor neutrophils to sepsis in addition to the immunosuppressive nature of sepsis neutrophils. We uncover the biology underpinning previously defined transcriptomic endotypes, in particular identifying expansion of *IL1R2+* immature neutrophils and enrichment of the response to IL-1 pathway in mature neutrophils in the poor outcome SRS1 state, consolidating the endotype status of the transcriptomic definitions. We validate these changes with integrative analysis of peripheral blood bulk RNA-seq data and single-cell CyTOF data from additional cohorts and trace immune dysfunction in sepsis through convalescent sample analysis to implicate a process of disrupted granulopoiesis.

Our data suggest that modulation of the IL-1 pathway is worth revisiting in biomarker-led sepsis clinical trials. While trials of IL-1 inhibition have previously failed to demonstrate mortality reduction(35, 36), these trials suffered from the absence of patient stratification. In a re-analysis of a phase III trial of IL-1R blockade with anakinra, improved survival was seen in a subgroup of anakinra-treated patients with hepatobiliary dysfunction and disseminated intravascular coagulation(37), suggesting the need for identification of the correct target patient group who may benefit from the therapy. Our data uncovering the basis of SRS endotypes suggest that there is potential utility in prospective trials of IL-1 inhibition in SRS1 patients specifically.

Our study also draws attention to the need for addressing the granulopoietic process in sepsis. Evidence for disrupted myelopoiesis in sepsis has previously been described in mice, including expansion of neutrophil progenitors at the expense of monocytes(38, 39). Previous clinical trials of G-CSF administration in sepsis have failed to show a reduction in mortality(40–42), while trials of GM-CSF are ongoing (43). However, an improved understanding of the mechanism by which these drugs influence neutrophils and granulopoiesis will be key. The addition of myelopoietic stimulus might not enhance the functional capacity of circulating immature and immunosuppressive cells, and the effects of such a stimulus on any already dysfunctional myelopoietic system are unknown and could even exacerbate the immunosuppressive actions of neutrophils in sepsis. Given the increased levels of G-CSF in SRS1 patients, G-CSF inhibition in SRS1 patients represents a potential alternative immunotherapeutic strategy.

In conclusion, our work provides insights into the innate immunobiology of sepsis, highlighting potential targets and pitfalls as we move towards molecular taxonomy-based approaches in personalized medicine for sepsis and other critical illness.

## Data Availability

All data produced in the present study are available upon request and will be made publicly available upon publication.

## Acknowledgments

We thank all the patients who participated in this study together with the GAinS investigators and recruiting hospitals. We acknowledge the support of the National Institute for Health Research through the Comprehensive Clinical Research Network for patient recruitment. This research was funded by the Medical Research Council (MR/V002503/1) (J.C.K. E.D.), Wellcome Trust Investigator Award (204969/Z/16/Z) (J.C.K), Wellcome Trust Grants (090532/Z/09/Z and 203141/Z/16/Z) to core facilities Wellcome Centre for Human Genetics, Wellcome Trust core funding to the Wellcome Sanger Institute (Grant number WT206194), and NIHR Oxford Biomedical Research Centre (J.C.K.). For the purpose of Open Access, the author has applied a CC BY public copyright license to any Author Accepted Manuscript version arising from this submission.

## Supplementary figure legends

**Figure S1.**
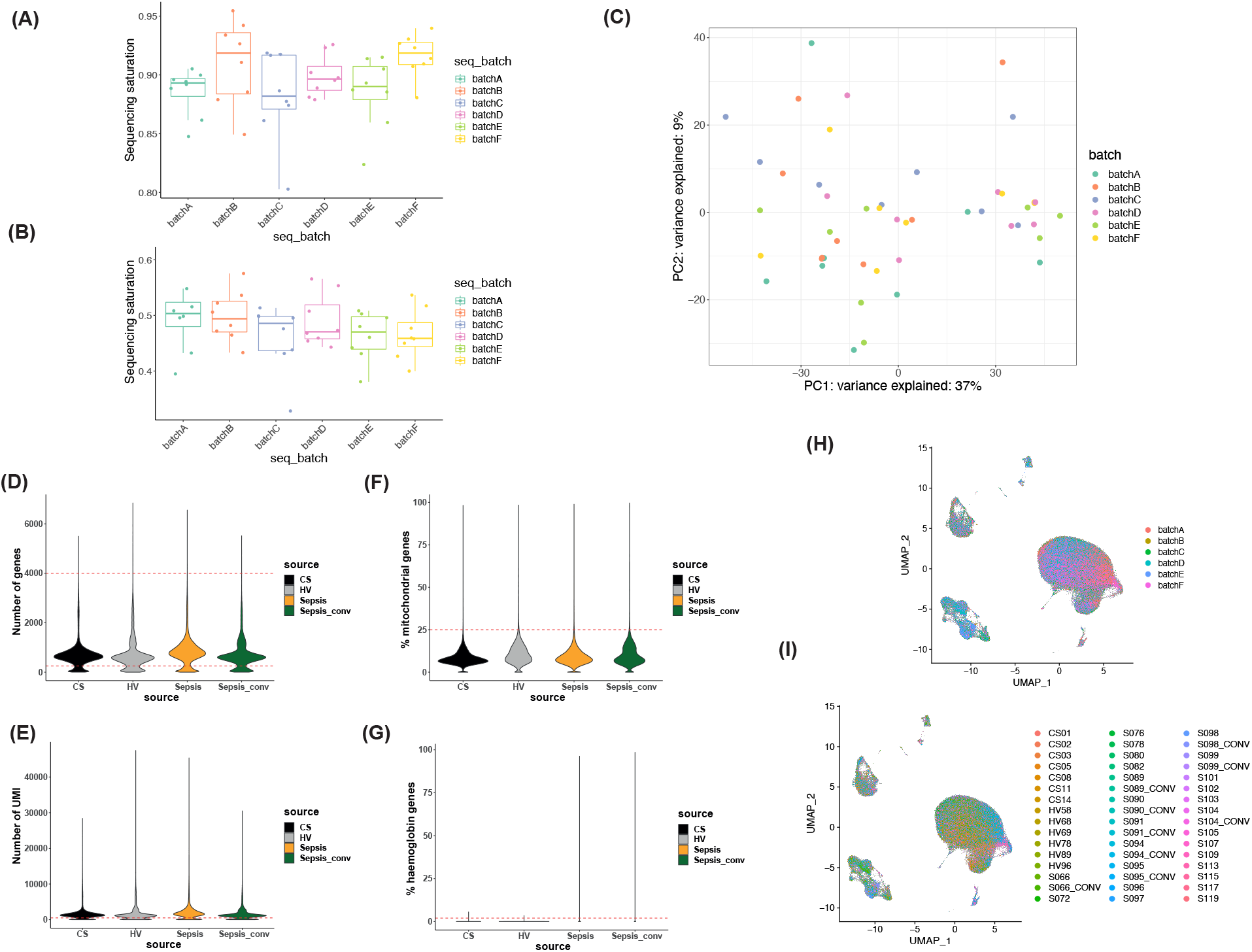
Single cell RNA and cell surface protein sequencing quality control (QC). (A-B) Sequencing saturation for single cell RNA (A) and protein (B) libraries per sequencing batch. (C) PCA of all 48 samples of cohort 1 colored by sequencing batch. (D-G) QC violin plots for various filtering thresholds (red dotted lines) for (D) number of genes detected per cell (E) number of UMIs per cell (F) percentage of mitochondrial genes detected per cell and (G) percentage of hemoglobin genes detected per cell. (H-I) UMAP visualisation of 272,993 cells after QC filtering colored by (H) sequencing batch or (I) sample identifier.

**Figure S2.**
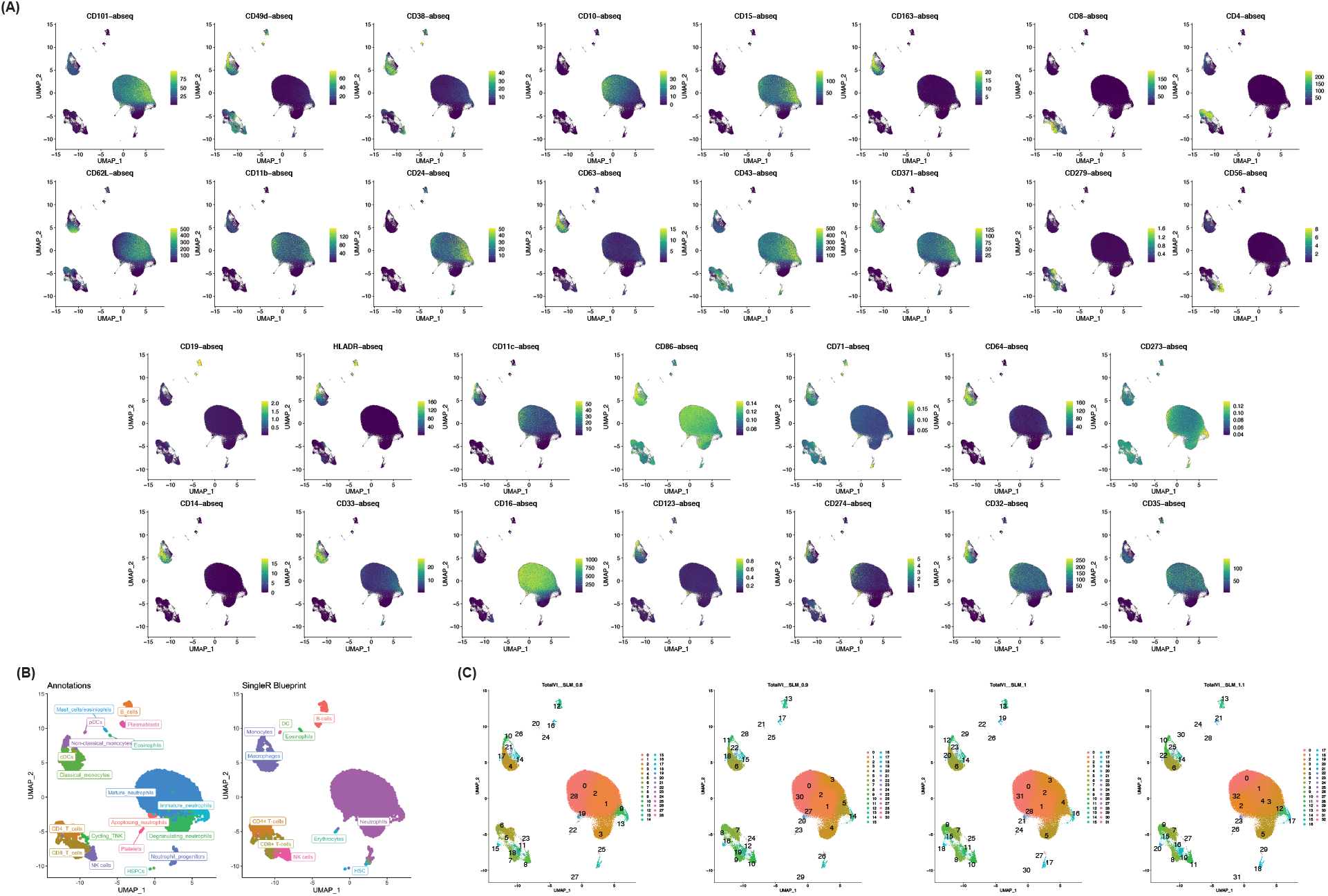
Annotation of and clustering single cell RNA and cell surface protein sequencing dataset. (A) UMAP visualisations of 272,993 cells with TotalVI denoised surface protein marker expression overlayed for 30 markers. (B) Comparison of broad manual annotations (“Annotations”) with data driven algorithmic labelling of cell state/identity by SingleR based on reference bulk RNA-seq profiles of pure cell populations from the Blueprint Consortium. (C) Clustering results at varying resolutions (0.8-1.1) with smart local moving algorithm.

**Figure S3.**
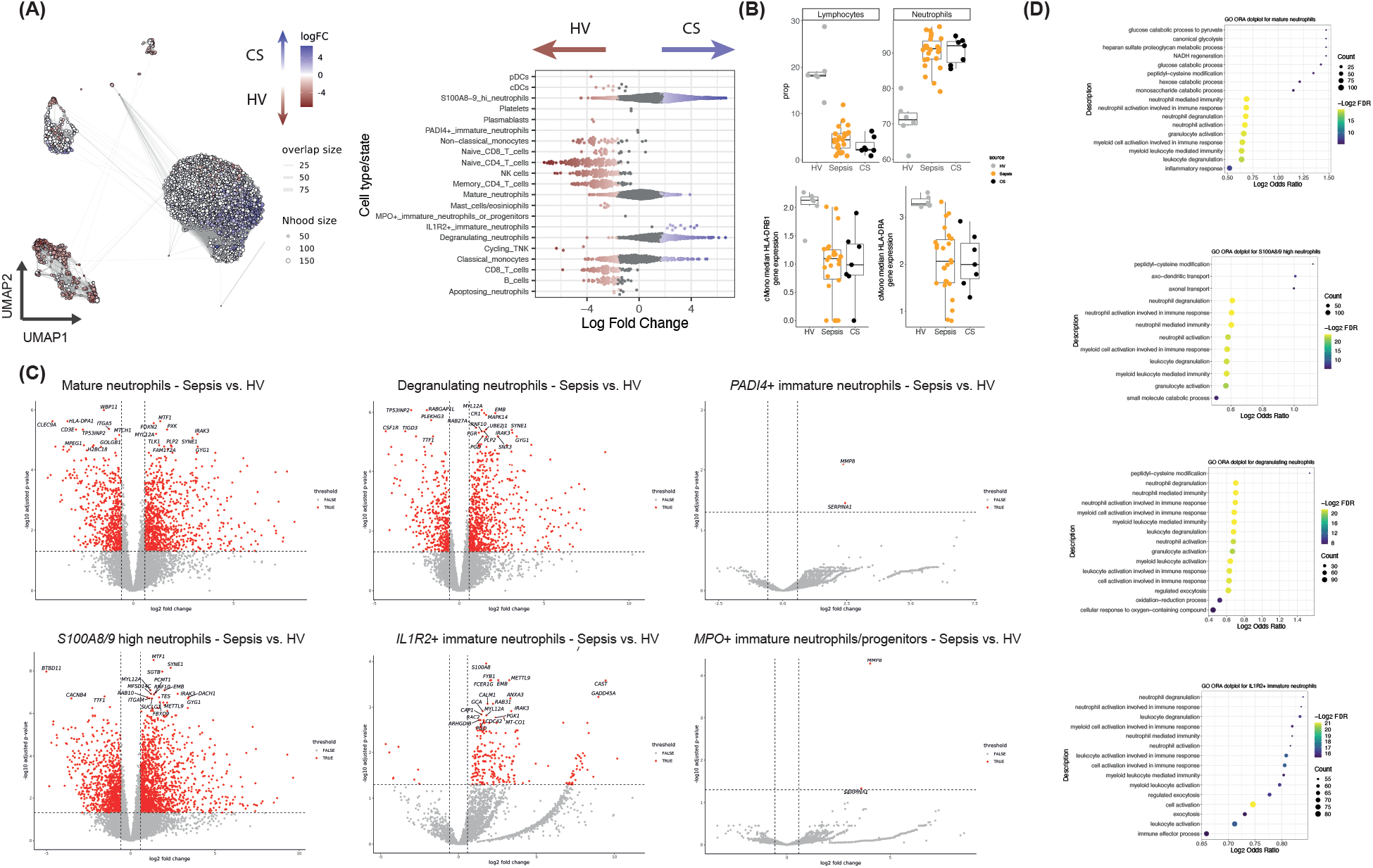
Neutrophil differential abundance (DA) and differential gene expression (DGE) analysis. (A) DA analysis using graph neighborhood-based method to detect enrichment of neighborhoods per comparator condition between CS and HV samples, with UMAP visualisation of sampled neighborhoods and statistically significant enrichment colored (spatial FDR < 0.05 with generalised linear modelling) (red = enrichment in HV, blue = enrichment in CS) and corresponding beeswarm plot (right) with cluster labels of neighborhoods depicted. (B) Boxplots of proportions of neutrophils and lymphocytes and *HLA-DRA* and *HLA-DRB1* gene expression in classical monocytes in HV, sepsis and CS samples. (C) Volcano plots of DGE analysis between sepsis and HV for pseudobulked mature, *S100A8/9* high, degranulating, *IL1R2*+ immature, *PADI4*+ immature and *MPO*+ immature/progenitor neutrophils (red denoting genes with fold change >1.5 and FDR < 0.05) (positive fold change denoting upregulation in sepsis). (D) Over-representation test pathway enrichment dotplots of top Gene Ontology (GO) pathways for genes upregulated in mature, *S100A8/9* high, degranulating and *IL1R2*+ immature neutrophils. *HV = healthy volunteers, CS = cardiac surgery patients, FDR = false discovery rate*.

**Figure S4.**
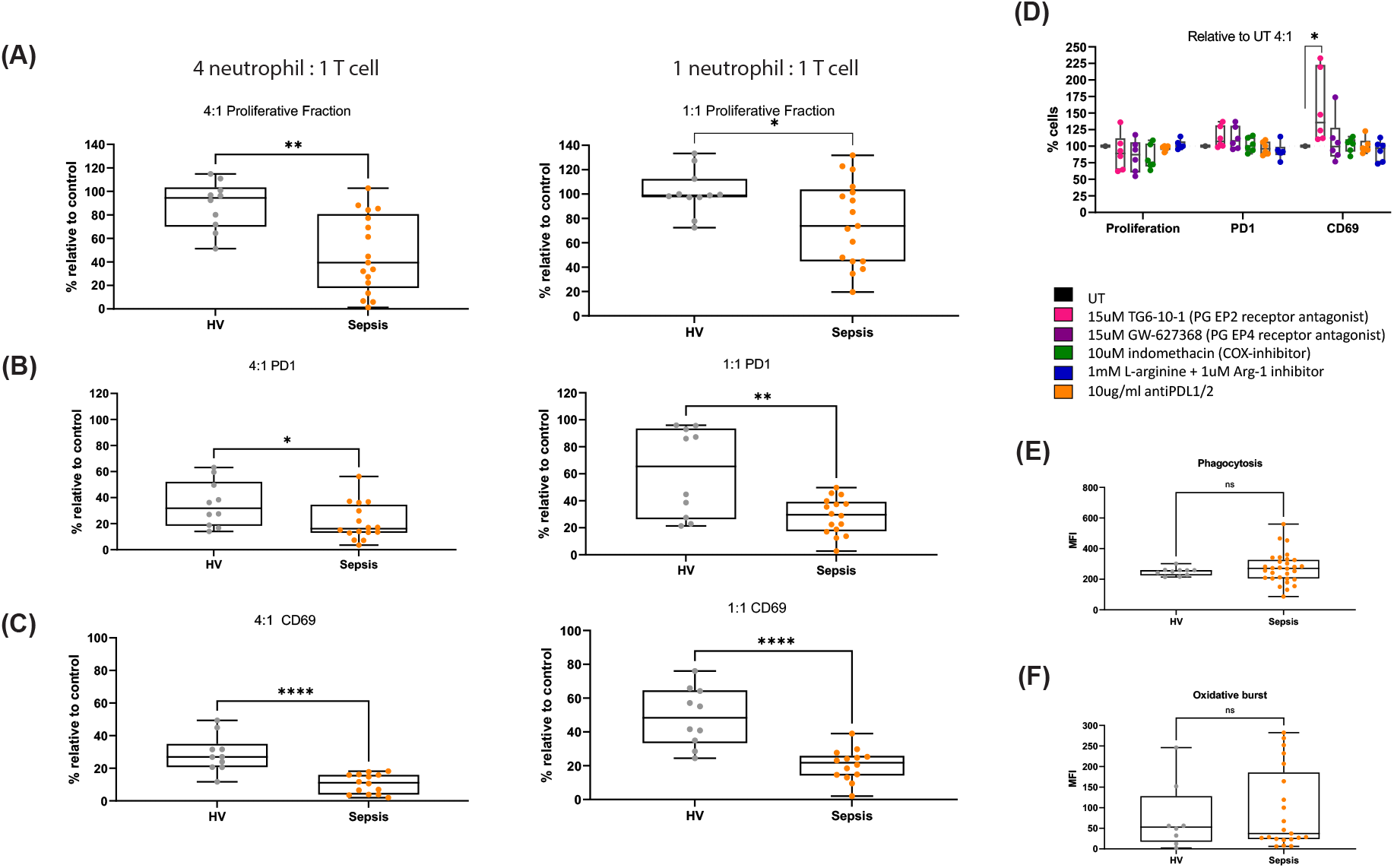
Sepsis neutrophil functional interrogation compared to HVs. (A-C) Boxplots of (A) proliferative fraction (B) surface PD-1 positive T cells and (C) surface CD69 positive T cells of anti-CD3/28 bead stimulated CD4 T cells after co-culture with neutrophils at a 4 neutrophil : 1 T cell ratio (left) and 1 neutrophil : 1 T cell ratio (right), compared to positive controls of CD4 T cells cultured with anti-CD3/28 beads alone. (D) Boxplots of percentage of CD4 T cells proliferating, expressing PD-1 or expressing CD69 in sepsis neutrophil-allogeneic T cell co-cultures with various treatments. (E) Boxplot of median fluorescence intensity (MFI) of neutrophils for phagocytosis assay with fluorescently labelled *E. Coli* conjugated phagocytosis beads. (F) Boxplot of median fluorescence intensity (MFI) of neutrophils for reactive oxygen species assay with dihydrorhodamine-123 incubation. *P*-values were calculated with two-sided Wilcoxon rank-sum tests. \**P* < 0.05, \*\**P* < 0.01, \*\*\**P* < 0.001. *UT = untreated, PG = prostaglandin, COX = cyclo-oxygenase*.

**Figure S5.**
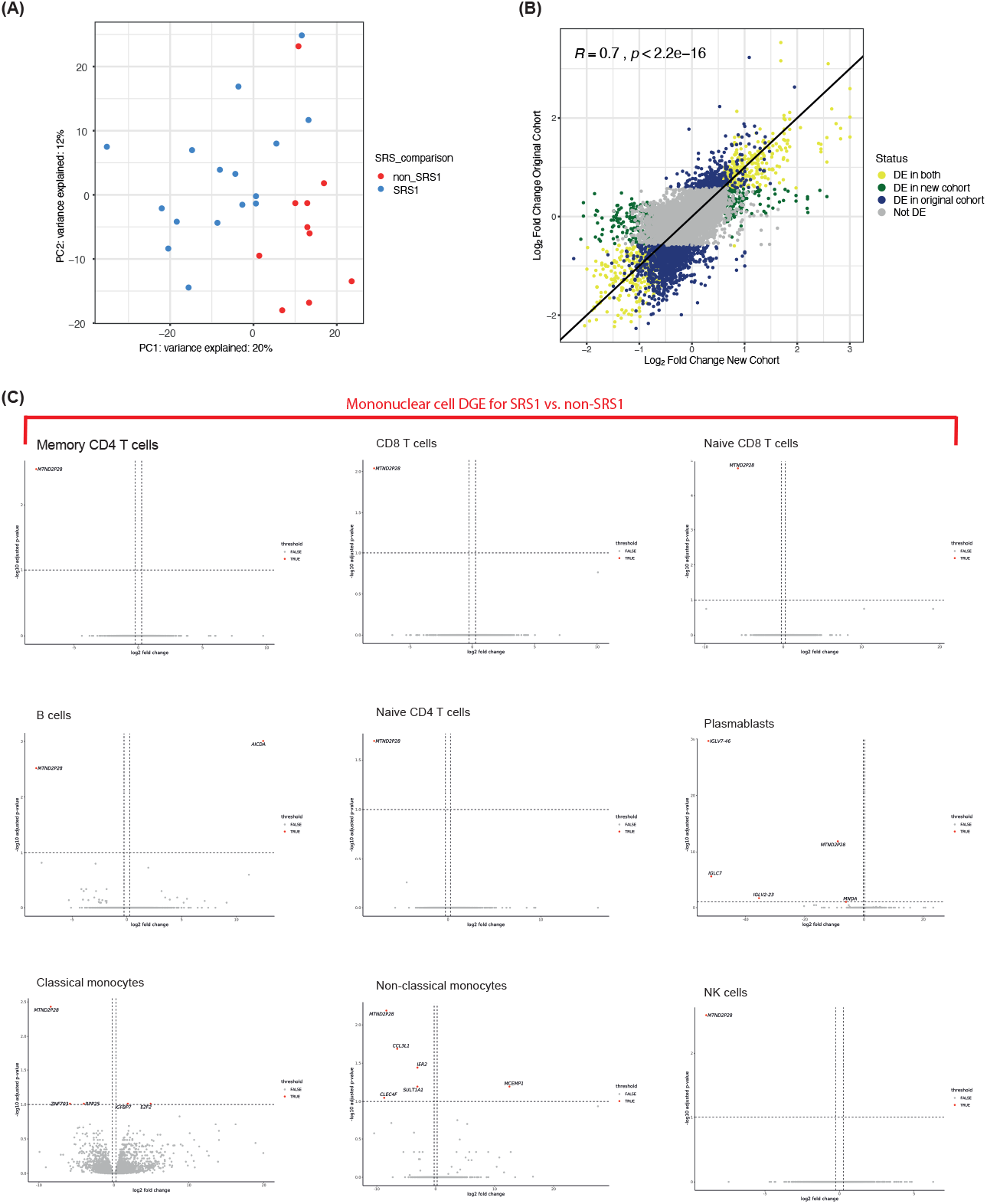
SRS transcriptomic endotype analysis and mononuclear cell pseudobulk SRS differential gene expression (DGE). (A) First two principal components from principal components analysis of pseudobulked single cell expression profiles of sepsis samples (all cells per sample) colored by SRS assignment. (B) Correlation of fold change in gene expression between SRS1 vs. non-SRS1 in cohort 1 and cohort in which SRS was originally derived. (C) Volcano plots of DGE analysis between SRS groups for pseudobulked mononuclear cells (red denoting genes with fold change > 1.5 and FDR < 0.05) (positive fold change denoting upregulation in SRS1).

**Figure S6.**
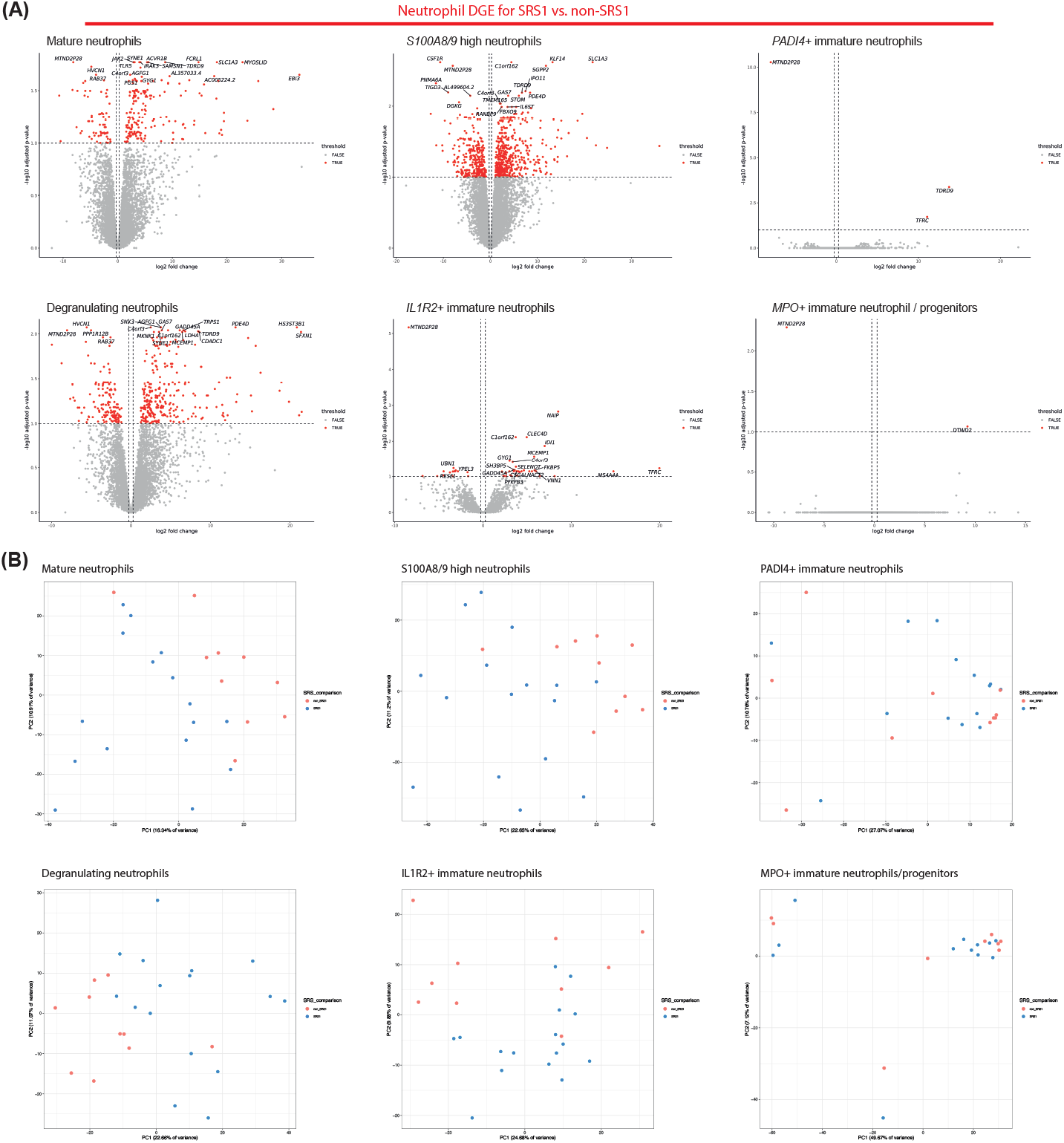
SRS neutrophil pseudobulk gene expression analyses. (A) Volcano plots of DGE analysis between SRS groups for pseudobulked neutrophil states (red denoting genes with fold change > 1.5 and FDR < 0.05) (positive fold change denoting upregulation in SRS1). (B) First two principal components (PCs) from PCAs of pseudobulked neutrophil states colored by SRS groups.

**Figure S7.**
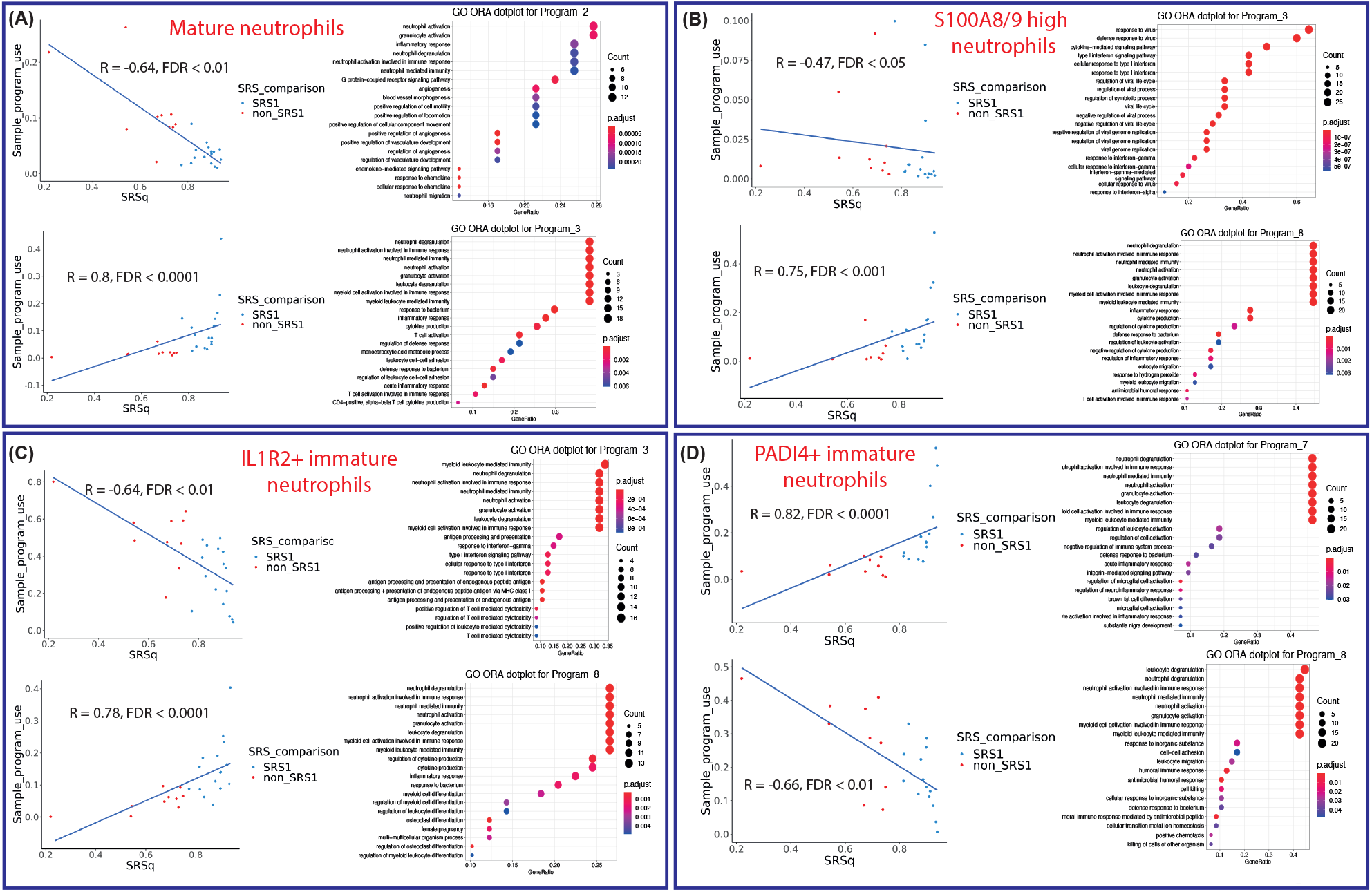
SRS neutrophil cNMF analyses. (A-D) Correlation plots of gene programs with program expression significantly correlated (Spearman *R* FDR < 0.05) with SRSq with corresponding over-representation test pathway enrichment dotplots of top Gene Ontology (GO) pathways for (A) mature neutrophils (B) *S100A8/9* high neutrophils (C) *IL1R2+* immature neutrophils and (D) *PADI4+* immature neutrophils.

**Figure S8.**
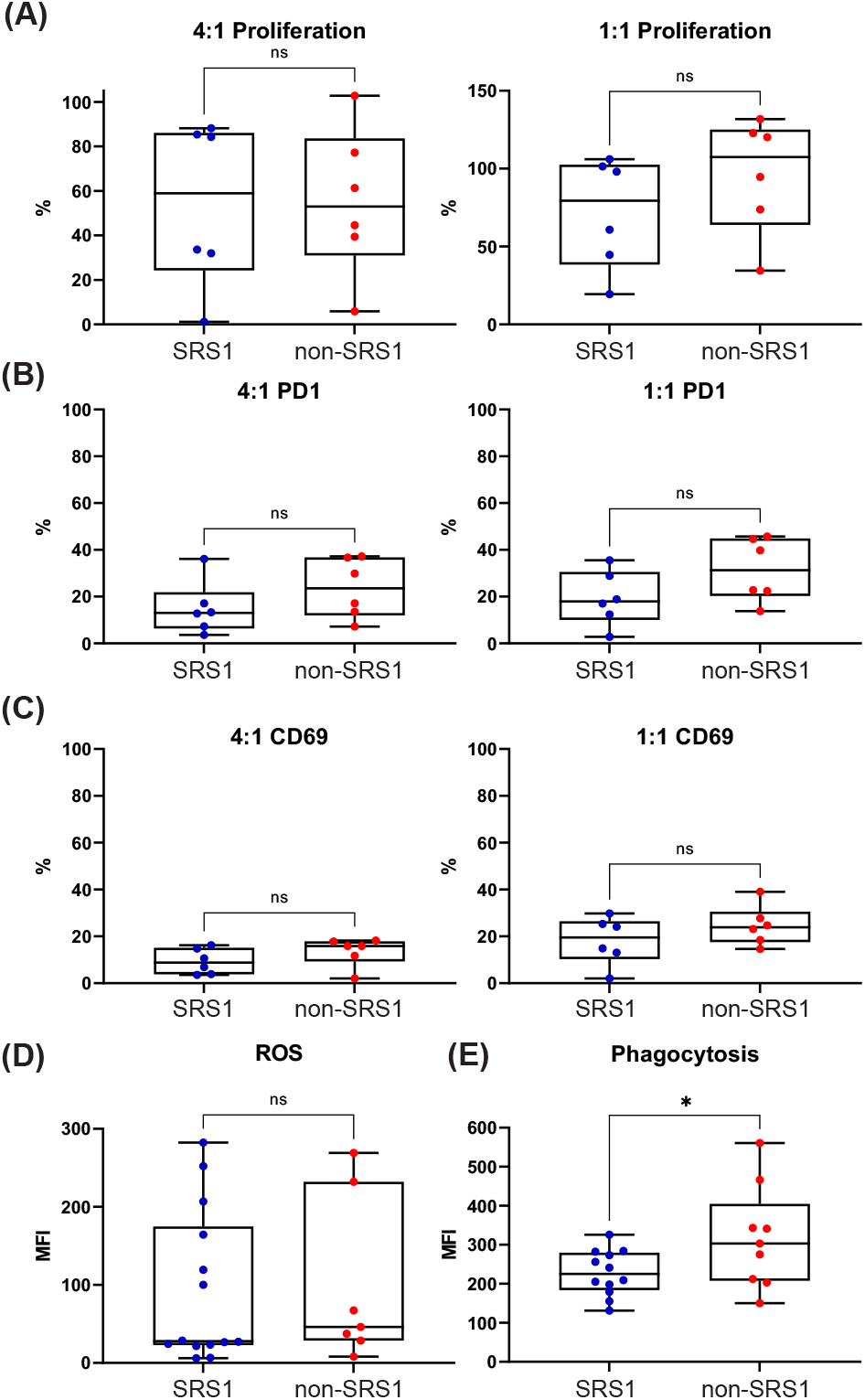
SRS neutrophil functional interrogation. (A-C) Boxplots of (A) proliferative fraction (B) surface PD-1 positive cells and (C) surface CD69 positive cells of anti-CD3/28 bead stimulated CD4 T cells after co-culture with neutrophils at a 4 neutrophil : 1 T cell (left) and 1 neutrophil : 1 T cell (right) ratios, compared to positive controls of CD4 T cells cultured with anti-CD3/28 beads alone. (D) Boxplot of median fluorescence intensity (MFI) of neutrophils for reactive oxygen species assay with dihydrorhodamine-123 incubation. (E) Boxplot of MFI of neutrophils for phagocytosis assay with fluorescently labelled *E. Coli* conjugated phagocytosis beads. *P*-values were calculated with two-sided Wilcoxon rank-sum tests. \**P* < 0.05.

**Figure S9.**
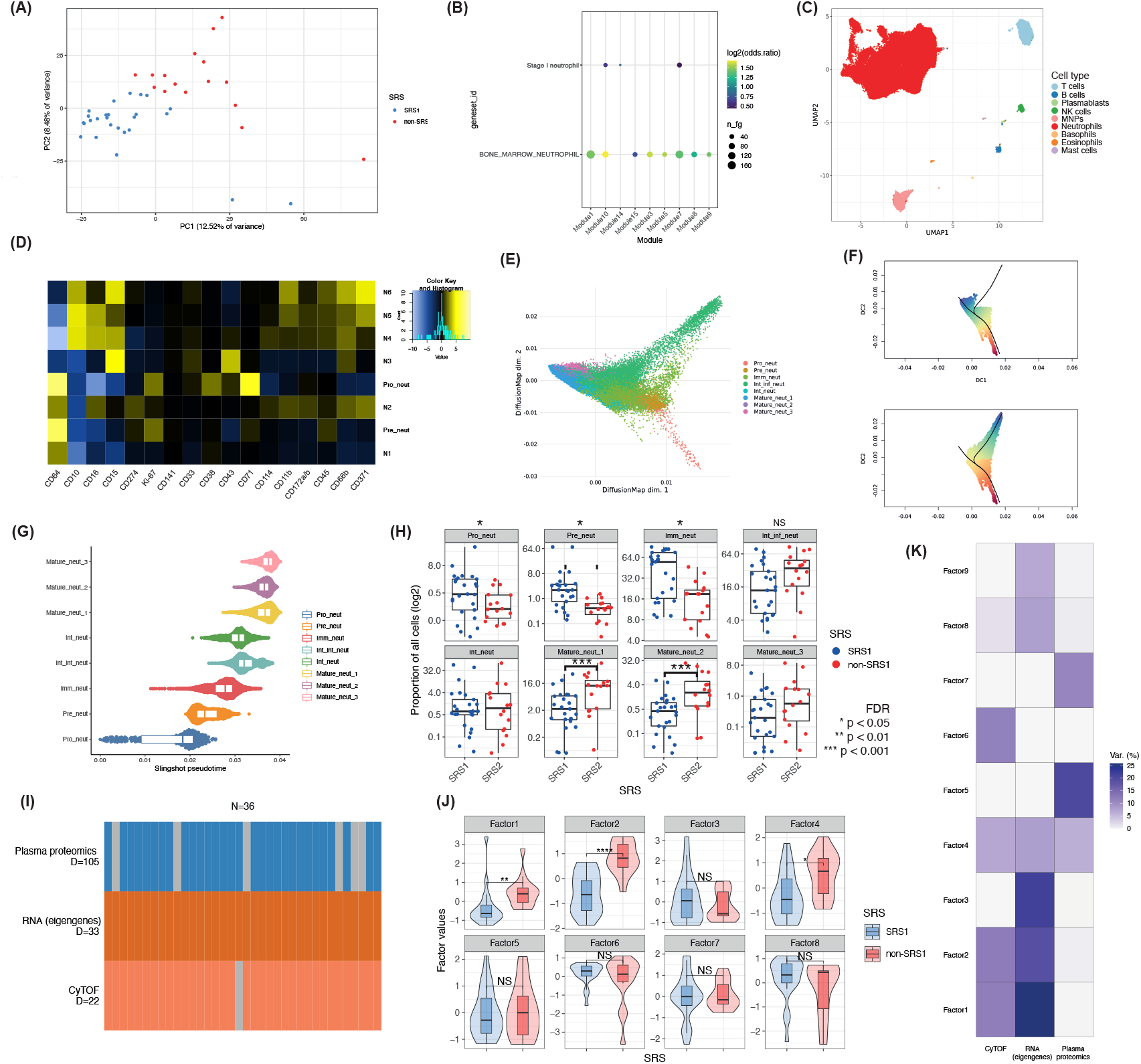
Cohort 2 SRS and CyTOF analysis and multi-modal factor analyses. (A) First two principal components (PCs) from PCA of n=42 sepsis patient samples (36 individuals) for peripheral blood bulk RNA-seq (23,063 genes) colored by SRS membership (3). (B) Enrichment of neutrophil related gene sets of WGCNA modules upregulated in SRS1 on overrepresentation testing against gene sets relating to neutrophil maturity. (C) UMAP visualisation of 9 broad immune cell types identified from self-organising map clustering (2,287,410 cells) of whole blood mass cytometry immunophenotyping data on 41 protein markers for n=41 sepsis samples (36 individuals) and n=11 healthy donor samples. (D) Marker enrichment modelling score heatmap of eight neutrophil subsets (1,921,471 cells) on 17 selected protein markers identified after neutrophil subclustering. (E) Visualisation of diffusion map dimension reduction of neutrophils (down sampled to 45,000 cells i.e. 15,000 per comparator group). (F) Scatterplots of diffusion components (DCs) 1 and 2 on 45,000 downsampled cells after diffusion map dimension reduction. Black lines depict two trajectories identified by principal curve fitting via *Slingshot*. Color reflects pseudotime value of cell with red indicating early pseudotime and blue late pseudotime. (G) Dotplot of neutrophil subsets according to pseudotemporal ordering from principal curve trajectory. (H) Boxplots of neutrophil cluster cell frequency as a proportion of all cells comparing SRS1 vs. non-SRS1 with proportional differences analysed by generalised linear mixed models (n=41 sepsis samples, 36 individuals). (I) Input data into MOFA+ model consisting of three modalities (105 plasma proteins, 33 module eigengenes from WGCNA and 22 cell types from CyTOF) for n=36 sepsis samples with one missing sample for CyTOF and six missing samples for plasma proteomics. (J) Violin and boxplots of factor values by SRS groups for each of eight latent factors identified with Wilcoxon rank sum testing. (K) Grid plot of variance decomposition showing percentage variance explained for individual modalities internally by each latent factor. FDR = false discovery rate, *FDR < 0.05, **FDR < 0.01, *** FDR < 0.001, **** FDR < 0.0001.

**Figure S10.**
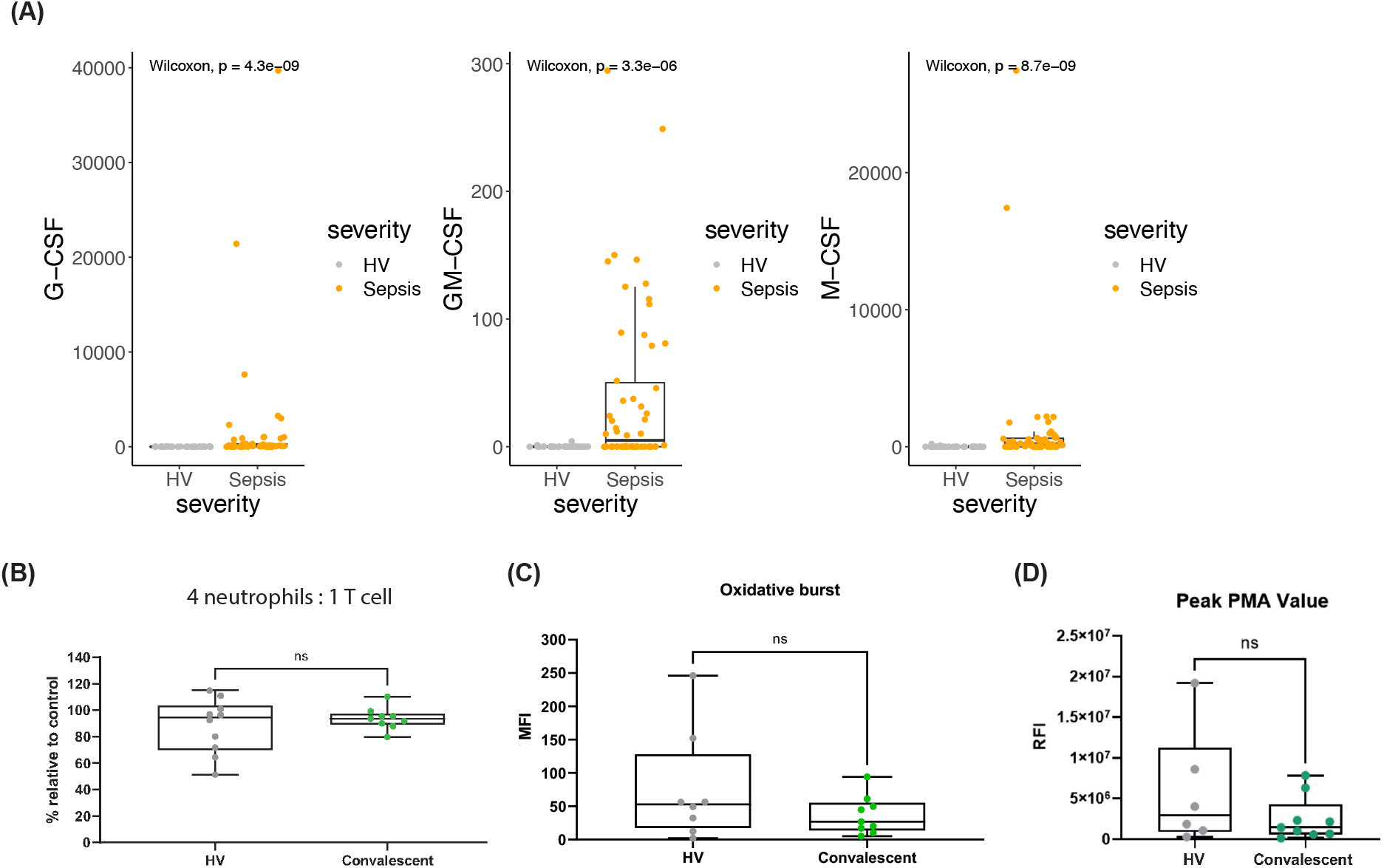
Cohort 2 cytokine analysis and cohort 1 convalescent neutrophil functional analyses. (A) Boxplots of plasma myelopoietic cytokine quantities between sepsis and HV groups in cohort 2. (B) Boxplots of proliferative fraction of anti-CD3/28 bead stimulated CD4 T cells after co-culture with neutrophils at a 4 neutrophil : 1 T cell ratio, compared to positive controls of CD4 T cells cultured with anti-CD3/28 beads alone. (C) Boxplot of median fluorescence intensity (MFI) of neutrophils for reactive oxygen species assay with dihydrorhodamine-123 incubation. (D) Boxplot of peak Cytotox green relative fluorescence intensity (RFI) per sample for HV and convalescent sepsis samples. *P*-values were calculated with two-sided Wilcoxon rank-sum tests.

## Methods

### Study ethics and patient cohorts

#### Cohort 1

Volunteers self-reporting as healthy and with no history of infection in the past 14 days were consented and recruited into the Genetic diversity and gene expression in white blood cells study [South Central Oxford REC B, reference 06/Q1605/55]. Blood samples were collected on one occasion.

Sepsis patient samples were collected from patients older than 18 years of age admitted to Oxford University Hospitals NHS Foundation Trust, UK. Patients were recruited either from the intensive care unit (ICU) if they had symptoms and signs of established sepsis (suspected infection with an acute change in total Sequential Organ Failure Assessment (SOFA) score of ≥2 points), or from the Emergency Department and medical wards if they had a change in quick SOFA score by ≥2 points and a NEWS2 score of ≥7 or intensive care review requested. Exclusion criteria included: patients or consultees unwilling or unable to give consent or advice declaration; advanced directive to withhold or withdraw life sustaining treatment; admission for palliative care only; pregnancy and 6 weeks post-partum; or severe acquired immunodeficiency including systemic high dose steroid therapy (prednisolone 0.5mg/kg/day for 14 days or equivalent), HIV infection, known regular therapy with immunosuppressive agents e.g. azathioprine, or neutrophil counts <1000/mL due to any cause including metastatic disease and haematological malignancies or chemotherapy but excluding severe sepsis and solid organ/bone marrow transplant recipient receiving immunosuppressive therapy.

Post cardiac surgery samples were collected from patients older than 18 years of age admitted to Oxford University Hospitals NHS Foundation Trust, UK. Patients were deemed eligible if they were 1) undergoing cardiac bypass surgery, 2) required post-operative intensive care unit stay and 3) did not have an infection prior to surgery. Exclusion criteria were identical to those for sepsis patients.

Sepsis and post cardiac surgery patients were consented into the Sepsis Immunomics Study [South Central Oxford REC C, reference:19/SC/0296] between May and November 2021. Samples were collected on days 1, 3 or 5 of either hospital or intensive care admission. Written informed consent was obtained from adults or advice declarations from personal/nominated consultees for patients lacking capacity, with retrospective consent obtained from the patient once capacity was regained.

#### Cohort 2

Healthy volunteers and sepsis patients were recruited into the same studies as in cohort 1 under the same criteria and ethics and were analysed previously as part of a COVID-19 centric bioresource (26).

The primary endpoint for the SRS groups was chosen to be 14-day mortality from sampling based on previous SRS analyses that early mortality was significantly different between the SRS groups(3, 10).

#### Cohort 3

Adult patients (≥18 years old) were recruited through the UK Genomic Advances in Sepsis (GAinS) study (NCT00121196) upon admission to intensive care with sepsis due to community-acquired pneumonia (CAP) or faecal peritonitis (FP); https://ukccggains.com/). Patients were recruited from 34 UK intensive care units (ICUs), with diagnoses, inclusion and exclusion criteria as described by Davenport et al(3). Ethics approval was granted nationally and locally (RECs 05/MRE00/38, 08/H0505/78, and 06/Q1605/55), with informed consent obtained from all patients or their legal representative.

### Fresh whole blood sample processing for single-cell sequencing

Blood samples were drawn with EDTA Vacutainer tubes (BD Biosciences) and processed within 1□h of collection. 1ml of whole blood was lysed with 9ml of 1X eBioscience red blood cell (RBC) lysis buffer (Thermofisher) for 10 minutes, quenched with 35 mL of Hanks’ Balanced Salt Solution (Lonza) and centrifuged at 300g for 10 minutes. The RBC lysis was repeated with 3 mL lysis buffer for 3 minutes and quenched and centrifuged again. The cells were then counted and 200,000 cells were transferred for antibody staining and single cell capture.

### Single cell RNA and cell surface protein profiling

Single cell RNA and cell surface protein sequencing was performed with the BD Rhapsody platform (Cat. No. 633731 and 633733), using the whole transcriptome amplification assay (Cat. No. 633801) and 30 Rhapsody Abseq antibodies (panel in Table S2). 1 uL of each Abseq antibody was added to 200,000 total white cells and topped up with 45 uL of BD staining buffer for 30 minutes. Samples were washed three times in 100 uL BD Rhapsody sample buffer + 200 uL BD staining buffer at 400g (5 minute centrifugation), before counting and diluting for single cell capture.

The concentration of cells was diluted to target a capture of 6000 cells per sample before loading onto the BD Rhapsody cartridge. Reverse transcription, complementary DNA (cDNA) amplification (Cat. No. 633773) and library construction (Cat. No. 633801) were performed following manufacturers’ recommendations. Libraries were sequenced to >80% saturation on NovaSeq S4 flowcells (Illumina).

### Single cell multi-omic analysis

#### Preprocessing and quality control

Gene expression data were aligned to the GRCh38 reference genome using STARsolo(44) (version 2.7.9a) with settings:

--soloType CB_UMI_Complex \

--soloUMIlen 8 \

--soloCBposition 0_0_0_8 0_21_0_29 0_43_0_51 \

--soloUMIposition 0_52_0_59 \

--soloFeatures Gene Velocyto \

--soloMultiMappers EM \

--soloCBmatchWLtype 1MM \

Unfiltered files i.e. containing all 884,736 cell barcodes on the whitelist were used downstream for an additional round of cell calling by the emptyDrops function within DropletUtils (v1.10.3) with a UMI threshold of 100 and FDR of 0.5%. Spliced and unspliced counts matrices were also produced with the Velocyto option in –soloFeatures.

Abseq reads were trimmed with Trimmomatic (v0.39) to 48 bases i.e. 12bp UMI + 36bp Abseq nucleotide sequence. The reads were then aligned to an artificial reference of the Abseq nucleotide sequences also using STARsolo with settings:

--alignIntronMax 1 \

--alignMatesGapMax 1 \

--winAnchorMultimapNmax 1 \

--chimMainSegmentMultNmax 1 \

--outFilterMultimapNmax 1 \

--seedMultimapNmax 1 \

Cells expressing fewer than 100 genes, more than 4000 genes, >10% mitochondrial reads, >2% haemoglobin reads and with a log10(UMI/gene) <0.6 were removed. Gene filtering was then performed with genes expressed in <10 cells or a total count of <3 removed.

Scrublet (v.0.2.3) and doublet detect (v3.0) were then both used on default settings to generate doublet scores. Automatic thresholds were applied to remove multiplets.

#### Normalisation, dimensionality reduction, visualisation and clustering

Data were normalised (scanpy v1.7.2: normalize_total), log□+□1 corrected (scanpy: log1p) and 4000 HVGs identified using the Seurat vst algorithm (scanpy: highly_variable_genes, flavor=‘seurat_v3’).

Multi-modal dimensionality reduction (DR) was performed with TotalVI (scvi-tools v0.10.0)(45) on default settings with all 30 proteins and 4000 HVGs with each individual sample set as a batch (as all samples were processed separately). The 20 latent dimensions identified were used for downstream UMAP visualisation and graph construction for clustering and differential abundance (DA) testing.

Unsupervised clustering was performed by first building a shared nearest neighbours graph with k=30 nearest neighbours in Seurat (v4.0). Choice of k parameter for number of nearest neighbours was made following inspection of Clustree (v0.4.20) analysis of a range of values for k and the effect on cluster splitting. Graph building was followed by clustering with the smart local moving algorithm in the Seurat (v4.0) FindClusters function (algorithm = 3).

Evaluation of clustering resolution was done by combining three metrics including assessment of cluster neighbourhood purity, cluster average silhouette width and a 30 iteration bootstrap to determine cluster stability with respect to sampling noise (bluster v1.0), and one subjective element i.e. inspection of clusters and their top defining markers to match clustering results with known biology and understand any potential value in merging vs. splitting. Combining these 4 elements, the default clustering resolution of 0.8 was chosen, with the splitting of one immature neutrophil cluster into two (MPO+ immature neutrophils/progenitors and PADI4+ immature neutrophils), based on the resolution of 1.1.

#### Cell type and state annotation and RNA velocity analysis

Cell types and states as defined at the above chosen resolutions and then merged for annotation at a broad level, and kept at the clustering resolution of choice for annotation at a fine granularity. Protein markers enabled identification of major lineages which were merged for the broad level annotations. Correct lineage assignment was confirmed by corroborating marker driven annotation with SingleR (v1.6.1) automatic assignments. Fine annotation was then performed based on inspection of biologically known/potentially meaningful gene markers to subdivide T cell and neutrophil populations.

The unspliced and spliced counts from STARsolo were used for RNA velocity analysis of the neutrophils (without degranulating or apoptosing neutrophils) with scVelo (v0.2.3). Moments were estimated using TotalVI reduced dimensions. The stochastic model was used for all analysis.

#### Differential abundance analysis

The 20 latent dimensions identified via multimodal DR with TotalVI(45) were used MiloR (v0.99.19)(19) kNN graph construction (k=30). The proportion of cells sampled for neighbourhood indexing was 0.1. Differential abundance (DA) testing was then performed with generalised linear models including age and sex as covariates. Significantly different neighbourhoods were determined by a spatial corrected FDR < 0.05.

#### Differential gene expression (DGE) analysis

For pseudobulk analysis, gene expression was aggregated per individual and per cell type/state into a pseudobulk count matrix i.e. raw UMI counts per gene were summed across all cells belonging to the same cluster and to the same individual. Genes were filtered per pseudobulk based on minimum expression of *n* counts in at least *X* samples, where *X* was the smallest sample group by source, and *n* was defined for each pseudobulk based on histograms of logged count distributions. Sample and cell type/state based pseudobulks were then normalised by the EdgeR trimmed mean of M-values (TMM) method (v3.30.3). The normalised count matrix was log transformed for principal components analysis (PCA).

Pseudobulk differential expression analysis was performed with a generalised linear model as implemented in EdgeR (v3.30.3), with the contrasts of interest specified with age, sex and sequencing batch included in the model.

Consensus DGE for convalescent vs. HV neutrophils was performed as described Wilk et al.(46). DEGs were taken as those with an average logFC > 0.25 (adjusted *P* < 0.05). Genes were filtered for those that showed reproducible change in the same direction in a minimum of 2/3 of samples i.e. 6 convalescent samples.

#### Consensus non-negative matrix factorisation (cNMF) and co-expression analysis

2000 highly variable genes for each neutrophil state were selected with the SelectIngerationFeatures function in Seurat (v4.0). Each of the neutrophil states were then subjected to cNMF as previously described (v1.2). Mean program usage per sample for each neutrophil state was correlated with SRSq scores per sample. The top 50 genes per program were taken for downstream pathway analysis.

To understand the genes linked to *TDRD9*, we calculated the Pearson correlation coefficient between the log normalised expression values of *TDRD9* and all other genes, and took the top 0.5% correlated genes for pathway analysis.

#### *IL1R2*+ neutrophil defining gene set

A gene set specific to *IL1R2+* neutrophils was identified by taking the *IL1R2+* neutrophil data from sepsis samples and looking for gene markers of these cells against all other cells with the FindMarkers function in Seurat (v4.0). The resulting markers were then filtered for an adjusted *P*-value of < 0.01 and an average log2FC >1 to obtain markers specific to the cells.

#### Gene set enrichment scoring for single cells

Gene sets for granulocytic myeloid derived suppressor cells (G-MDSC)(21, 22) were scored in the dataset using the AddModuleScore function in Seurat (v4.0), followed by Wilcoxon rank sum tests to test for differences across different groups of cells.

#### Pathway enrichment analysis

Pathway enrichment analysis was performed against Gene Ontology Biological Process pathways via the ClusterProfiler R package (v4.1.4) (47) with over-representation analysis via Fisher’s exact tests as implemented in the enrichGO and enricher functions.

#### Sepsis endotype (sepsis response signature) assignment/scoring

To assign SRS endotypes for sepsis samples, gene expression was aggregated per individual for all cells to produce a per sample pseudobulk count matrix i.e. raw UMI counts per gene were summed across all cells belonging to the same individual. Genes were filtered then per pseudobulk based on minimum expression of 5 counts in at least *X* samples, where *X* was the smallest sample group by comparator group i.e. six HVs. Sample pseudobulks were then normalised by the EdgeR trimmed mean of M-values (TMM) method (v3.30.3). The relevant genes (*DYRK2, CCNB1IP1, TDRD9, ZAP70, ARL14EP, MDC1*, and *ADGRE3*) from the normalised count matrix were then used for SRS assignment and SRSq score calculation by SepstratifieR (v0.0.0.9)(23).

### Isolation of neutrophils from whole blood

CD66b+ cells were isolated from 5-10ml whole blood from EDTA Vacutainer tubes (BD Biosciences) using EasySep HLA Chimerism Whole Blood CD66b positive selection kit (StemCell) following manufacturer’s instructions. Cells were washed with HBSS (Lonza) and resuspended in complete media (RPMI-1640 10% FCS, 100 IU/ml L-glutamine, 100 IU/ml penicillin-streptomycin) ready for downstream assays.

### Oxidative Burst and Phagocytosis Assays

Neutrophils were incubated in 5ml polypropylene test tubes for 20 minutes in complete media with or without pHrodo Green E.Coli Bioparticles (Invitrogen) at a concentration of 1 in 10 for assessing phagocytosis or with DihydrorhodamineR-123 (Invitrogen) at a final concentration of 7.5uM for assessing oxidative burst. Cells were washed and stained for surface markers for 30 minutes with 7AAD, CD66b-AF700 (G10F5) and Siglec 8-APC (7C9) antibodies from Biolegend and fixed using Fix/Lyse solution (Invitrogen) for 10 minutes before acquisition using BD LSRFortessa X-20 analyser. The median fluorescent intensity (MFI) for single 7AAD^-^ CD66b^+^ Siglec 8^-^ gated cells was determined by subtraction of the MFI of the *fluorescence minus one* (FMO) control sample that was not incubated with any pHrodo bioparticles or Dihydrorhodamine-123.

### NETosis assay

Neutrophils were plated at 20,000 cells/well in low auto-fluorescence F-12K Nut Mix (Gibco) media (Hams-F12) using flat-bottomed 96-well plates treated for 1hr with 0.01% poly-L-ornithine solution (Sigma). Cells were allowed to attach for up to 1hr prior to treatment with 100nM PMA, as per Incucyte assay recommendations, or left untreated as controls. All cells were also treated with 250nM IncuCyte Cytotox Green Dye, used to measure NETosis-induced cell death. The final volume in each well was made up to 100ul using Hams-F12 media. The plated cells were then transferred to the IncuCyte Live-Cell Analysis System incubator and scans were scheduled for every 20mins for up to 24 hours, using standard scan type, objective 20X with green fluorescence channel selection with vessel type 96-well plate.

### Neutrophil - allogeneic CD4 T cell co-culture

Cryopreserved CD4+ T cells isolated from healthy donor leukocyte cones were thawed and stained with eBioscience Cell Proliferation Dye eFluor450 (Invitrogen) at a final concentration of 10uM for 20 minutes following manufacturer’s recommendations. T cells were then immediately cocultured with the isolated neutrophils in 96 well U bottom plates at 37°C 5% CO_2_ at either a 4:1 or 1:1 neutrophil to T cell ratio in complete media supplemented with 50 IU/ml recombinant human IL-2 (Biolegend) and with anti-CD3/CD28 dynabeads (Gibco). The volume of anti-CD3/CD28 beads added was equivalent to a 1:1 bead to T cell ratio and the total cell density was maintained at 200,000 cells per well. As controls, T cells were also plated without anti-CD3/CD28 beads and with anti-CD3/CD28 beads only i.e. no coculture, for every run to model resting and activated levels respectively. Some cells were also incubated with 1mM L-arginine (Sigma-Aldrich), 1uM Arg-1 inhibitor CB-1158 (Fisher Scientific), anti-CD274 (eBioscience clone MIH1), anti-CD273 (PD-L2) (eBioscience clone MIH18), 20ug/ml PGE_2_ (Sigma-Aldrich), 10uM indomethacin (Sigma-Aldrich), 15uM EP_2_ inhibitor TG-6-10-1 (MedChemExpress) or 15uM EP_4_ inhibitor GW627368 (MedChemExpress).

After 3-4 days of coculture, cells were stained for surface markers for 30 minutes at room temperature followed by staining with Annexin-V and 7AAD for 15 minutes to determine the percentage of live non-apoptosing cells for each sample. Samples were analysed using BD LSRFortessa X-20. All antibodies were purchased from Biolegend unless otherwise stated: CD3-APC (UCHT1), CD4-BUV395 (BD Biosciences, SK3), CD66b-AF700 (G10F5), PD1-PE (NAT105), CD69-PECy7 (FN50), 7AAD, Annexin V-FITC. T cells were gated from a live single cell population as CD66b^-^ CD4^+^ CD3^+^ Annexin-V^-^ 7AAD^-^ cells. Proliferation analysis by dye dilution was established using the naive T cells as the baseline for determining the proliferative fraction of cells in each sample. The proliferative fraction and percentage of cells expressing the markers PD-1 and CD69 are displayed as a percentage relative to that of the anti-CD3/28 bead stimulated no coculture T cells control to account for donor variation.

### Flow cytometry and NETosis data analysis

All flow cytometry data was analysed using Flowjo V.10. Figures and statistics were created using Graphpad Prism v9. Details of statistical analyses performed are included in all figure legends.

### Cell type/state deconvolution of 864 sepsis bulk RNA-seq samples

Bulk RNA sequencing data were obtained for 864 samples (667 patients) from the GAinS study. Briefly, whole blood samples were taken on the first, third, and/or fifth day following ICU admission and leukocytes were isolated using the LeukoLOCK system (Thermo Scientific). RNA was extracted using the manufacturer’s protocol (Ambion), and libraries prepared using NEB Ultra II Library Prep kits (Illumina) were sequenced on a NovaSeq 6000 (Illumina). Reads were aligned to the GRCh38 reference genome using STAR (v2.7.3) and gene expression quantified using featureCounts. Finally, the count matrix was normalised using TMM from edgeR and log-transformed.

A signature matrix for CIBERSORTx was constructed using our single cell dataset, with each finely annotated cell type and state except apoptosing cells downsampled to 100 cells per type/state. The signature matrix was then created via the Create Signature Matrix analysis module with a parameters min. expression = 0.25, replicates = 100 and sampling = 0.5.

Cell fractions of the 864 sample whole blood bulk RNA-seq dataset were then imputed with the Impute Cell Fractions analysis module using the single cell reference matrix, with batch correction S-mode enabled and quantile normalisation disabled.

### RNA-seq exploratory analysis, weighted gene co-expression network analysis (WGCNA) and pathway analyses for cohort 2

The edgeR trimmed mean of M-value normalised(48) and filtered count matrix was obtained from the COMBAT consortium(26), with 23,063 features from 42 samples analysed.

Principal components analysis was performed with all features transformed at log2(counts per million + 1). PCA was carried out using prcomp (R v4.0.0) with default parameters, while hierarchical clustering was done on Euclidean distance with Ward’s method as implemented in hclust (R v4.0.0).

WGCNA was conducted as per the vignettes from (https://horvath.genetics.ucla.edu/html/CoexpressionNetwork/Rpackages/WGCNA/Tutorials/) with the log normalised count matrix. The soft thresholding power of 4 was first chosen by taking the lowest power which reached a scale free topology model fit of 0.8. This was then used to construct a signed-hybrid network with biweight midcorrelation and a deesplit parameter of 2 with a minimum module size of 30. Modules were summarised by eigengene expression (the value of the first principal component of the module) and modules with eigengene correlation of <0.2 were merged. Differential testing of modular eigengene expression across SRS groups was conducted with Wilcoxon rank sum tests, adjusting for multiple testing with the Benjamini-Hochberg method.

Pathway enrichment analysis was performed against Gene Ontology Biological Process pathways and custom gene sets via the ClusterProfiler R package(47) with over representation analysis via Fisher’s exact tests as implemented in the enrichGO and enricher functions. Gene set enrichment analysis for *IL1R2+* neutrophil defining genes for the WGCNA module most correlated with immature neutrophil proportions was performed via the GSEA function.

### CyTOF pre-processing, clustering, trajectory inference and differential abundance analyses for cohort 2

Normalised, debarcoded, and bead and doublet cleaned FCS files were obtained from the COMBAT consortium(26). To avoid biases due to highly varying cell numbers per sample, a maximum of 50,000 cells per sample was taken. As not all batches contained control samples for batch correction, a control sample free correction method *Harmony(49)* was adapted for use given previous demonstration of utility in CyTOF data(50).

The batch corrected matrix was then fed into clustering analysis via FlowSOM(51) as implemented in the CATALYST package(52). This was first done with all cells to a resolution of 144 clusters (12×12 starting grid) with metacluster merging by consensus clustering to 50 metaclusters, and following manual annotation, performed again by subsetting the neutrophil clusters and reclustering to 30 metaclusters. Neutrophil clusters were manually annotated based on median marker expression and then compared with marker enrichment modelling scores for analytical validation. The number of cells identified in each cluster (including neutrophil subclusters) was divided by the total number of cells in the patient to evaluate the proportion of the cell type/state.

Neutrophils were downsampled to 15,000 cells per comparator group (SRS1, non-SRS1 and healthy volunteers) for diffusion map dimension reduction as implemented in the CATALYST package(52) followed by trajectory analysis with the Slingshot package*(30)*. A principal curve was fitted on the minimum spanning tree with default settings on the slingshot function.

Principal components analysis (PCA) was performed after quantile normalising the cell proportions of all 22 cell types/states. PCA was carried out using prcomp (R v4.0.0) with default parameters.

Differential abundance analysis was performed using a generalised linear mixed model to account for repeated sampling in six patients, with batch included as a covariate in the model and patient included as a random effect. An extra random effect term for each individual sample was included to model the overdispersion in proportions seen in high dimensional cytometry data(52). Adjustment for multiple testing of each cell type was performed with the Benjamini-Hochberg method.

### Multi-modal analysis by MOFA+ for cohort 2

Concentrations of 105 plasma proteins as determined by the timsTOF mass spectrometer were obtained from the COMBAT consortium(26).

For each sample, the eigengenes of the 33 identified modules from WGCNA, quantile normalised proportion of the 22 cell types from CyTOF clustering and annotation, and log2 transformed and median centered intensity values for 105 plasma proteins (as in the previous analysis) (26) were utilised for factor analysis as implemented in the MOFA+ R package(31). Views were left unscaled, number of factors was set at 10, and all other model options were left with default values. Training convergence was done in slow mode and other options were left with default values. Differences in factor values between SRS groups was tested with a Wilcoxon rank-sum test, adjusting for multiple testing with the Benjamini-Hochberg method.

### Cytokine analysis for cohort 2

Concentrations of 51 plasma analytes as determined by the Luminex assay were obtained from the COMBAT consortium(26). Differential testing of cytokine concentrations across sepsis vs. healthy states or across SRS groups was conducted with two-sided Wilcoxon rank sum tests.

## Notes

### Competing Interest Statement

The authors have declared no competing interest.

### Author Declarations

Ethical approval for this work was given by the following UK Research Ethics Committees: South Central Oxford REC B (06/Q1605/55), South Central Oxford REC C (19/SC/0296), Scotland A Research Ethics Committee (05/MRE00/38), and Berkshire Research Ethics Committee (08/H0505/78).

